# Protocol for a sequential, prospective meta-analysis to describe coronavirus disease 2019 (COVID-19) in the pregnancy and postpartum periods

**DOI:** 10.1101/2020.11.08.20228056

**Authors:** Emily R. Smith, Erin Oakley, Siran He, Rebecca Zavala, Kacey Ferguson, Lior Miller, Gargi Wable Grandner, Ibukun-Oluwa Omolade Abejirinde, Yalda Afshar, Homa Ahmadzia, Grace Aldrovandi, Victor Akelo, Beth A. Tippett Barr, Elisa Bevilacqua, Justin S. Brandt, Natalie Broutet, Irene Fernández-Buhigas, Jorge Carrillo, Rebecca Clifton, Jeanne Conry, Erich Cosmi, Camille Delgado-López, Hema Divakar, Amanda J. Driscoll, Guillaume Favre, Valerie Flaherman, Christopher Gale, Maria M. Gil, Christine Godwin, Sami Gottlieb, Olivia Hernandez Bellolio, Edna Kara, Sammy Khagayi, Caron Rahn Kim, Marian Knight, Karen Kotloff, Antonio Lanzone, Kirsty Le Doare, Christoph Lees, Ethan Litman, Erica M. Lokken, Valentina Laurita Longo, Laura A. Magee, Raigam Jafet Martinez-Portilla, Elizabeth McClure, Torri D. Metz, Deborah Money, Edward Mullins, Jean B. Nachega, Alice Panchaud, Rebecca Playle, Liona C. Poon, Daniel Raiten, Lesley Regan, Gordon Rukundo, Jose Sanin-Blair, Marleen Temmerman, Anna Thorson, Soe Soe Thwin, Jorge E. Tolosa, Julia Townson, Miguel Valencia-Prado, Silvia Visentin, Peter von Dadelszen, Kristina Adams Waldorf, Clare Whitehead, Huixia Yang, Kristian Thorlund, James M. Tielsch

## Abstract

We urgently need answers to basic epidemiological questions regarding SARS-CoV-2 infection in pregnant and postpartum women and its effect on their newborns. While many national registries, health facilities, and research groups are collecting relevant data, we need a collaborative and methodologically rigorous approach to better combine these data and address knowledge gaps, especially those related to rare outcomes. We propose that using a sequential, prospective meta-analysis (PMA) is the best approach to generate data for policy- and practice-oriented guidelines. As the pandemic evolves, additional studies identified retrospectively by the steering committee or through living systematic reviews will be invited to participate in this PMA. Investigators can contribute to the PMA by either submitting individual patient data or running standardized code to generate aggregate data estimates. For the primary analysis, we will pool data using two-stage meta-analysis methods. The meta-analyses will be updated as additional data accrue in each contributing study and as additional studies meet study-specific time or data accrual thresholds for sharing. At the time of publication, investigators of 25 studies, including more than 76,000 pregnancies, in 41 countries had agreed to share data for this analysis. Among the included studies, 12 have a contemporaneous comparison group of pregnancies without COVID-19, and four studies include a comparison group of non-pregnant women of reproductive age with COVID-19. Protocols and updates will be maintained publicly. Results will be shared with key stakeholders, including the World Health Organization (WHO) Maternal, Newborn, Child, and Adolescent Health (MNCAH) Research Working Group. Data contributors will share results with local stakeholders. Scientific publications will be published in open-access journals on an ongoing basis.

## Introduction

Severe acute respiratory syndrome coronavirus 2 (SARS-CoV-2) has led to over 244 million confirmed cases of coronavirus disease 2019 (COVID-19) and claimed more than 4.9 million lives globally, as of October 2021 [1], and these are likely underestimates [2]. The World Health Organization (WHO) and the U.S. Centers for Disease Control and Prevention (CDC) note that pregnant women may be at higher risk of developing severe illness due to COVID-19 [3,4]. The specific mechanisms for this increased risk are unknown, though may be related to physiologic and immunologic changes during pregnancy. Pregnant women are generally at increased risk for severe illness from many infectious diseases including influenza, hepatitis E, malaria, and herpes simplex virus [5]. Additionally, other coronaviruses such as severe acute respiratory syndrome (SARS) are thought to pose higher risks for adverse outcomes to pregnant people; although there is only a single case-control study comparing 10 pregnant to 40 non-pregnant SARS patients [6,7].

Pregnant women with COVID-19, and their offspring, may be at increased risk for complications due to severe illness and adverse pregnancy outcomes. A living systematic review suggests that pregnant women with COVID-19, compared to non-pregnant women with COVID-19, are more than twice as likely to be admitted to an intensive care unit (ICU) or require invasive ventilation [8]. In an observational cohort study of 1,219 pregnant women across 33 U.S. hospitals with positive SARS-CoV-2 test (molecular or antigen) at the time of delivery, severe or critical COVID-19 was associated with a 40% increase in hypertensive disorders of pregnancy and a 42% increase in preterm birth, as compared to asymptomatic pregnant patients [9]. A multi-national case-control study (INTERCOVID) including 706 pregnant women with COVID-19 and 1,424 pregnant women without COVID-19 found that COVID-19 diagnosis was associated with increased risk of preeclampsia and eclampsia, severe infections, intensive care unit (ICU) admission and death [10]. A pooled analysis of 41 studies including 3323 pregnant women with COVID-19 found that one-third of their newborns were admitted to the neonatal intensive care unit (NICU); a meta-analysis of ten studies found that newborns born to pregnant women with COVID-19 were nearly five times more likely to be admitted to the NICU as compared to COVID-19 negative pregnancies [8]. Reports of mother-to-child transmission of SARS-CoV-2 during the antepartum, intrapartum, or early postnatal period remain limited.

Despite the urgent need to accurately document the number of cases, severe illness, and deaths in pregnant women, available data on maternal and infant complications related to COVID-19 remain heterogenous [11]. The high degree of heterogeneity in study designs, comparison groups, and definitions of key health outcomes across studies makes it difficult to clearly understand any excess risks and guide policy for pregnant and postpartum women and their newborns. For example, the current (February 2021) living systematic review on COVID-19 in pregnancy included 192 studies with 64,676 pregnant and postpartum patients; the heterogeneity in published outcomes meant that only a small fraction of those studies could be combined for key analyses, and data from some regions of the world, (e.g., sub-Saharan Africa) were absent. For example, data were available from only a handful of studies to examine a few risk factors for a single outcome (severe COVID and death), and there were fewer than 10 studies that compared the risk of maternal, fetal, and newborn outcomes between pregnant and postpartum patients versus either pregnant patients without COVID-19 or nonpregnant women of reproductive age with COVID-19 [8]. Further, we have limited data on the efficacy of treatments and preventives in pregnancy and postpartum, due largely to the systematic exclusions of pregnant people from the relevant clinical trials until recently. The same was true of the seminal vaccine trials, although vaccine studies among pregnant women began in early 2021 (ClinicalTrials.gov Identifier: NCT04754594) [12]. Establishing mother-to-child transmission of SARS-CoV-2 has also been challenging because consensus was lacking on case definition for intrauterine, intrapartum or early postpartum transmission of SARS-CoV-2 until February 2021 [13], and all proposed definitions have required clinicians and researchers to follow specific testing protocols [14–18].

COVID-19 data in pregnancy and infancy was scarce at first, and now it is heterogeneous. These shortcomings exist in part because data collection protocols have differed, and the underlying population risks for severe illnesses vary globally. It is therefore necessary to generate high-quality information with a unified data harmonization plan and analytic strategy. This approach will permit us to make evidence-driven decisions and to create guidelines for the prevention and treatment of COVID-19 in pregnant women. Because little of the existing data can be pooled, our ability to make strong recommendations related to COVID-19’s risks in pregnancy and the postpartum period is limited. We are similarly constrained in our capacity to offer guidance as to the potential benefits of new interventions, like vaccines. So we propose to fill this gap by conducting a sequential prospective meta-analysis (PMA). This PMA will be a robust, collaborative, global approach to accrue harmonized data, with the objective of generating policy and practice-relevant data about the epidemiology of COVID-19 in the pregnancy and postnatal periods.

## Methods

The protocol for this serial prospective meta-analysis was registered with PROSPERO (ID: 188955) on May 28, 2020. A prospective meta-analysis identifies studies that will contribute data to the meta-analysis, as well as establishes the analysis plans, before the results of the individual studies are known [19]. This approach is similar to a multi-site registry or cohort in the sense that studies work to harmonize collection of key outcomes, but differs from multi-site studies in that each site implements a study design and local protocol that is appropriate for their context [19]. Our goal is to provide a maximally flexible, robust, collaborative, and inclusive analysis framework that is methodologically appropriate.

### Ethical Approval

All individual study sites will obtain appropriate ethical approval for their independent data collection activities. As this project is a meta-analysis using aggregate study estimates or analyses of secondary, de-identified individual patient data, it is not classified as human research and thus is exempt from IRB approval.

### Language

Not all those who are pregnant or give birth identify as women. We use the terms “pregnant women” and “pregnant people” throughout. This accurately reflects both study designs (e.g. a comparison group including those who self-report as women between the ages to 15 and 45) and the language used by the studies contributing data.

### Research Questions

The study aims to answer basic epidemiological questions about COVID-19 and its impact on maternal and newborn health by pooling data from independent studies using harmonized data definitions and an individual patient data meta-analytic framework to minimize data variability. Analyses will be performed in three groups. First, we will provide descriptive statistics among pregnant women with COVID-19. To understand the relative risks associated with COVID-19 in pregnancy, we will compare pregnant people with COVID-19 to i) other pregnant people without COVID-19 and ii) non-pregnant women of reproductive age (15-45 years) with confirmed or suspected COVID-19.

Specific objectives regarding COVID-19 among pregnant or postpartum women include: i) describe the natural history of COVID-19; ii) estimate the incidence of and risk factors for hospitalization, admission to intensive care unit, receipt of critical care, and use of invasive ventilation (for COVID-19); and iii) estimate the infection and case fatality rate. For each of these outcomes, we will also compare the risk for pregnant women with COVID-19 to non-pregnant women of reproductive age with confirmed or suspected COVID-19.

Specific objectives regarding maternal health among pregnant or postpartum women with COVID-19 are to estimate the incidence or prevalence of: i) maternal morbidities including hypertensive disorders of pregnancy, abnormal placentation, preterm prelabor rupture of membranes, hemorrhage, embolic disease, etc.; ii) maternal and pregnancy-related death; iii) adverse pregnancy outcomes including stillbirth, preterm birth, small-for-gestational age birth, and low birthweight. For each of these outcomes, we will also compare the risk for pregnant women with COVID-19 to other pregnant women without COVID-19.

Specific objectives regarding newborn health among newborns born to patients with COVID-19 are to estimate the incidence and timing of and risk factors for: i) congenital anomalies; ii) perinatal and (early) neonatal mortality; and iii) intrauterine, intrapartum, or early peripartum transmission of SARS-CoV-2 from mother to child. When appropriate, we will compare these risks to those of newborns born to women without COVID-19 during pregnancy.

Specific objectives regarding SARS-CoV-2 in biospecimens (e.g. nasopharyngeal, vaginal, or rectal swab, maternal blood, pregnancy tissue, placenta, amniotic fluid, cord blood, breast milk) include estimating: i) the proportion of biospecimens with detectable SARS-CoV-2 virus and median viral load; ii) the association between virus or viral load in biospecimen and key outcomes described above.

### Search strategy & study inclusion

We originally recruited study sites to join the proposed prospective meta-analysis first via professional research networks, and subsequently via key stakeholder networks. Stakeholders at the National Institute of Child Health and Human Development (NICHD) at the U.S. National Institutes of Health (NIH) supported recruitment of NIH-funded maternal and child health network groups and other U.S. government funded projects. Stakeholders at the World Health Organization (WHO) in the Departments of Maternal, Newborn, Child, and Adolescent Health (MNCAH) and of Sexual and Reproductive Health and Research (SRH) supported recruitment of researchers engaged in the COVID-19 MNCAH research network and the SRH pregnancy cohorts working group [20]. Stakeholders from the International Federation of Gynecology and Obstetrics (FIGO) supported recruitment by issuing an invitation through their international network. Studies were invited to participate based solely on study design. Eligible study designs included: i) registries enrolling all suspected or confirmed cases in pregnancy or postpartum period, ii) cohorts enrolling all pregnant women, or iii) case-control studies enrolling pregnant or postpartum women with suspected or confirmed COVID-19. There were no *a priori* sample size limitations due to the dynamic epidemiology of the pandemic. Study investigators confirmed their intent to contribute to the PMA via a letter of intent or signing a collaboration agreement.

Building on concepts laid out in the Framework for Adaptive Meta-analyses (FAME) [21], we will also collaborate with the PregCOV-19 Living Systematic Review Consortium to identify studies that might be eligible for post-publication inclusion into the proposed meta-analysis. The search strategy for the LSR was previously published [8,22]. We will screen all published studies included in the living systematic review for potential inclusion in the PMA using the following criteria: i) the study conforms to the study designs outlined above; ii) there is a defined catchment area (e.g., certain hospitals, states, etc.); and iii) the sample size included at least 25 pregnant or postpartum people with confirmed or suspected COVID-19 who were consecutively recruited or identified through surveillance.

### Exposure (Suspected or Confirmed SARS-CoV-2 Infection)

Confirmed cases of COVID-19 will be defined as those with laboratory-confirmed SARS-CoV-2 infection via a nucleic acid amplification test, regardless of clinical signs or symptoms. The original protocol was expanded to include COVID-19 cases confirmed via antigen tests as they became validated and widely used over the course of the pandemic. Suspected cases will be defined according to the WHO August 7, 2020 case definition based on either a) clinical (acute onset fever and cough, or acute onset of three or more pre-identified signs and symptoms) and epidemiological criteria (residing, working, or traveling in an area with high risk of transmission or community transmission in the past 14 days or working in any health care setting) or the severe acute respiratory illness (SARI) case definition [23]. Probable COVID-19 infections will also be defined according to the WHO August 7, 2020 case definition (clinical criteria and contact with a case; a suspected case with chest imaging, recent anosmia or ageusia onset without other cause, respiratory disease preceding death and contact with a case) [23].

### Outcomes of Interest

Outcomes related to women’s health will include:

- Mortality: all-cause mortality, COVID-19 specific mortality, and pregnancy-related and maternal mortality;
- COVID-19 related clinical signs and symptoms: fever, cough, shortness of breath, dizziness or fainting, body aches, runny nose, sore throat, loss of sense of smell, loss of sense of taste, sneezing, fatigue, nausea, vomiting, diarrhea, and headache;
- Morbidity and healthcare utilization related to COVID-19 severity: pneumonia diagnosis, hospitalization, admittance to an intensive care unit (ICU), or use of critical care interventions;
- Pregnancy-related morbidities: hypertensive disorders of pregnancy including gestational hypertension, preeclampsia, eclampsia, hyperemesis gravidarum, fetal growth restriction, abnormal placentation (placenta previa and placenta accreta spectrum), placental abruption, preterm labor, preterm prelabor rupture of membranes, hemorrhage (antepartum/intrapartum; postpartum; abortion-related), embolic disease, or anesthetic complications. We also collected data on cesarean delivery.
- Adverse pregnancy outcomes: stillbirth (fetal death ≥28 weeks’ gestation per WHO definition), early preterm birth (<34 weeks gestation), preterm birth (<37 weeks gestation), small-for-gestational-age birth (<3rd and <10th percentile per the INTERGROWTH-21^st^ newborn reference values) [24], short-for-gestational-age birth (<3rd and <10th percentile per the INTERGROWTH-21^st^ newborn reference values on length [25], low birthweight (<2500 grams), and very low birthweight (<1500 grams).
- Presence of SARS-CoV-2 and viral load in maternal biological specimens including: amniotic fluid, placenta (maternal or fetal side), cord blood, vaginal swab, feces or rectal swab, nasopharyngeal swab, pregnancy tissue (fetus or pregnancy sac and placenta) in the case of stillbirth or abortion, breastmilk, and maternal blood.

Neonatal outcomes of interest include:

- Congenital anomalies: neural tube defects, microcephaly, congenital malformations of ear, congenital heart defects, orofacial clefts, congenital malformations of digestive system, congenital malformations of genital organs, abdominal wall defects, chromosomal abnormalities, reduction defects of upper and lower limbs, or talipes equinovarus (clubfoot);
- Early neonatal (≤7 days) and neonatal (≤28 days) death;
- Perinatal death (stillbirth (≥28 weeks) or early neonatal death (≤7 days);
- Admission to and length of stay in neonatal intensive care unit (NICU);
- Mother-to-child transmission of SARS-CoV-2 (differentiating intrauterine versus intrapartum or early peripartum infection if possible).

### Data Harmonization

We developed the draft data modules and questions in April 2020 based on a proposed set of questions from the Pregnancy CoRonavIrus Outcomes RegIsTrY (PRIORITY) study (ClinicalTrials.gov Identifier: NCT04323839). We also reviewed and included questions from the case report and data collection forms developed by the Department of Sexual and Reproductive Health (SRH), HRP, WHO and the WHE, WHO prospective cohort study, and the U.S. Centers for Disease Control and Prevention (CDC) Surveillance for Emerging Threats to Mothers and Babies Network (SET-NET) study [26,27]. We requested two rounds of feedback via a survey and by email from the >50 participants of the bi-monthly informal meeting established at the beginning of the pandemic called the “Perinatal COVID-19 Global Gathering”. The current data modules reflect feedback and general consensus among survey respondents. The final draft of the data modules and core variables was finalized and shared broadly on June 2, 2020 (S File 1). We updated the data modules in September 2020 to reflect evolving understanding of SARS-CoV-2 infection in newborns and to reflect the updated SRH, HRP, WHE UNITY generic protocol developed by WHO for COVID-19-related pregnancy cohort studies [26] (S File 2).

### Data Sharing & Study-Specific Data Analysis

We will develop a codebook and statistical codes for each contributing study to map original study variables to the PMA core variables. Investigators can join the PMA either by submitting individual patient data or running standardized code to generate aggregate data estimates that can be included in the meta-analysis. The same data quality and consistency checks will be performed for each study, and any issues will be resolved with study investigators. Studies will be eligible to contribute data to the PMA when they have accrued at least 25 confirmed cases with completed follow up.

Study-specific estimates for the two-stage meta-analysis will be produced using standardized analytical codes, and aggregated measures will be exported into a database for the meta-analyses. Individual studies are expected to proceed with their own study publications and can use any analyses generated by the PMA for publication or policy making in their study context.

### Overlapping Data

For each updated meta-analysis, we will review included studies for potentially overlapping participants to avoid including individual participants in the estimates for more than one study. We will choose a single study to contribute cases from given locations (e.g. health facilities, cities, states, or countries). The study with the most complete data will be selected to contribute, or in the case of similar data between studies, we will include data from the study that was the first to join the PMA. The number of studies and the number of cases per study is therefore likely to change with each updated meta-analysis, given this robust analytical strategy. We will use a modified version of the Newcastle Ottawa Scale to assess the risk of bias of individual studies.

### Methods for Data Synthesis

Ideally, all individual-level data would be combined for one-stage meta-analysis. However, we anticipate that the ability to quickly share data and the degree of willingness to share individual patient data may vary by country and across collaborators and institutions. Thus, we will plan for a step-wise statistical analysis plan where the most feasible and simple analyses (that contribute directly to our research questions) are prioritized, and more advanced statistical modelling will be conducted subsequently.

For the first stage of analysis, prevalence, and incidence data for the respective outcomes, overall and by risk factor, will be pooled using the conventional DerSimonian-Laird random-effects model [28]. For analyses where only proportions or crude incidence rates are used, the Arcsine method may be applied to stabilize the statistics and ensure approximate asymptotic normality [29]. For analyses comparing pregnant COVID-19 cases to a) COVID-19 negative pregnancies or b) non-pregnant women with COVID, we will calculate the relative risk or odds ratio (for case-control studies) for each outcome; effect estimates will be pooled using the conventional DerSimonian-Laird random-effects model [28]. We will assess forest plots visually for heterogeneity. When at least ten studies are being pooled, we will also quantify heterogeneity by the *I*^*2*^ statistic [30].

### Analysis of Subgroups

Where appropriate and as sample size allows, we will consider meta-regression or subgroup analyses by the following study level characteristics: study design and sampling strategy, proportion of confirmed COVID-19 cases (out of suspected and confirmed cases), national maternal mortality ratio, national neonatal mortality rate, and geographic region. We will also consider subgroup analysis by the following individual patient characteristics to identify risk factors or effect modifiers for specific outcomes: confirmed versus suspected COVID-19 case status; gestational age at COVID-19 onset (by week or by trimester), COVID-19 severity, pre-pregnancy health conditions or comorbidities [diabetes, hypertension, cardiovascular disease, obesity (Body Mass Index ≥30 kg/m^2^), underweight (Body Mass Index <18.5 kg/m^2^), tuberculosis, malaria, HIV/AIDS, syphilis, anemia (hemoglobin < 11 g/dL)], maternal morbidities (also described above as outcomes), maternal vaccination status (influenza, COVID-19), calendar time or time since first COVID-19 diagnosis in the study area, parity, maternal age, race or ethnicity, and maternal education. Some of these risk factors may be considered both outcomes of having COVID-19 in pregnancy or factors that exacerbate SARS-CoV-2 infection in pregnancy; we will attempt to disambiguate between the two based on the timing of diagnoses whenever possible.

### Sample size considerations

Consistent with the GRADE event-based approach, we will avoid “very low precision” by requiring a minimum number of three sites or a total of 100 events for each outcome to run the first analysis [31]. As data accumulate, we will evaluate the robustness of the inferences and whether the answer can be reasonably inferred via a conservative sample size. Assuming at least 50% heterogeneity between sites, the need for 90% power and 5% type 1 error to detect a difference, a conservative 1% prevalence/incidence and a risk ratio of 1.5 for the considered subgroup, a minimum of 30,000 participants are required to demonstrate an effect for each outcome and the proposed subgroups (risk factors). Alternatively, a total of 400 events or more may be used as a threshold for ‘sufficient evidence’ in accordance with recommendation of the GRADEPROfiler [31,32]. When these thresholds are met, the steering committee must decide whether additional, updated meta-analyses will be performed.

### Governance

The steering committee will consist of at least one member from each participating site, key stakeholders (S File 3), and the technical coordinating team at the George Washington University. When a formal vote is needed, the following teams will each cast one official vote: each group of investigators linked to participating studies, each key stakeholder organization, and the technical coordinating team. The steering committee will prioritize research questions and agree on common elements of data collection. They will disseminate results, including rapid reports to key stakeholders, webinars, and submission of manuscripts to preprint servers and scientific journals. The technical coordinating team will develop protocols for data transfer and ensure data quality; write the statistical analysis plans; and conduct meta-analyses.

## Participating studies

At the time of publication, 25 studies (in 41 countries) were actively participating in the ongoing PMA (Table 1). Among these studies, 11 have a contemporaneous comparison group of pregnancies without COVID-19; two studies include a comparison group of non-pregnant women of reproductive age. The median anticipated sample size of participating studies is 1,500. More than 76,000 pregnancies in total are expected to contribute to the completed meta-analyses (Table 1). These studies include data from 41 countries: Argentina, Australia, Bangladesh, Belgium, Brazil, Canada, Chile, China, Colombia, Democratic Republic of the Congo, Egypt, France, French Guyana, The Gambia, Germany, Ghana, Guatemala, Hong Kong, India, Israel, Italy, Kenya, Lebanon, Mali, Mexico, Mozambique, Netherlands, New Zealand, Nigeria, Pakistan, Peru, Portugal, Puerto Rico, Rwanda, South Africa, Spain, Sweden, Switzerland, Turkey, Uganda, United Kingdom, United States, and Zambia (Fig 1). Data from additional countries is expected from the WHO prospective cohort study and other studies that will be identified via the published literature. A description of each study, including the study investigators and institutional affiliations, study design description, recruitment methods, anticipated sample sizes, recruitment timeline, and primary outcomes are presented in supplementary tables (S3 File).

**Table 1.**
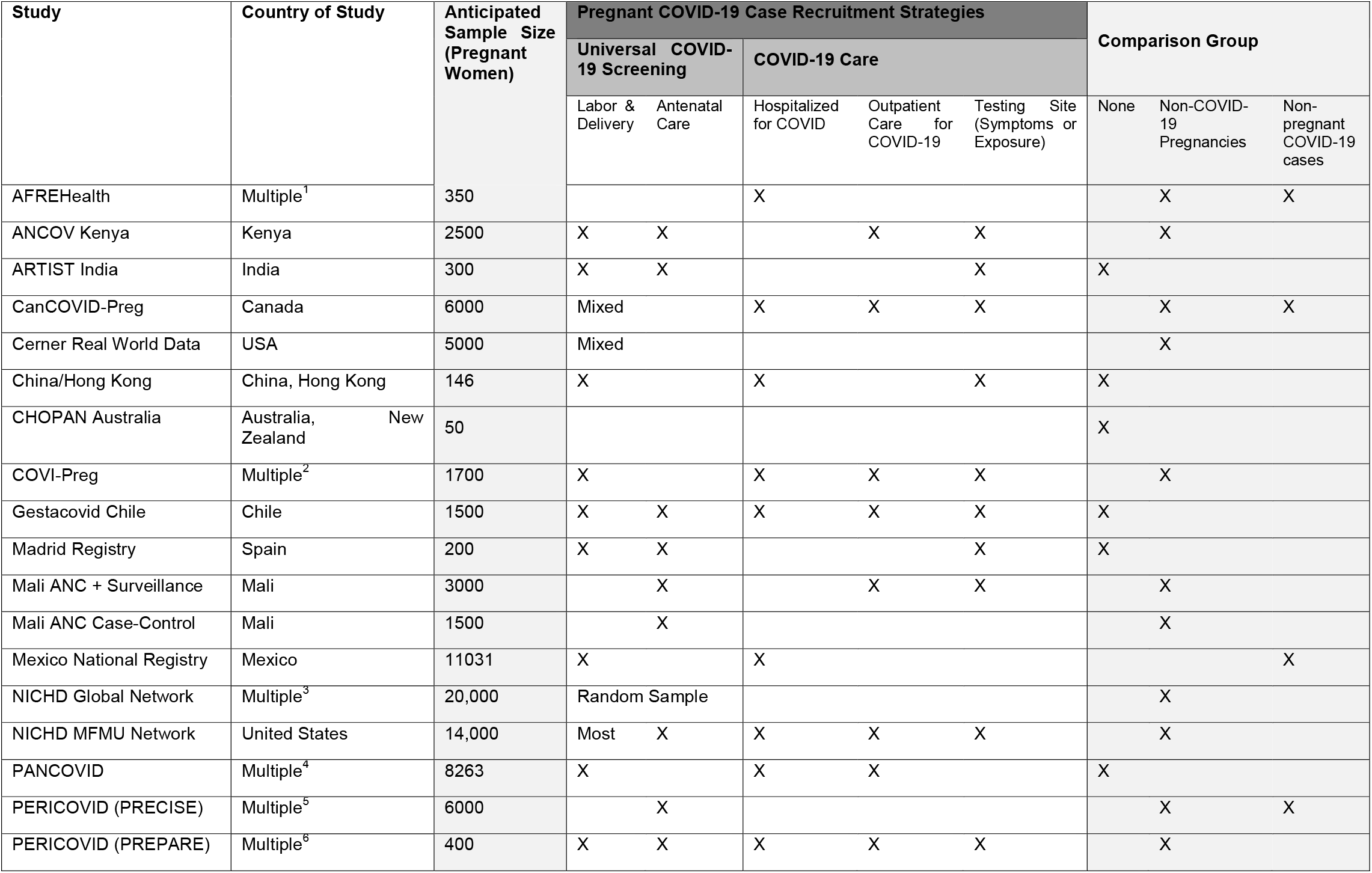
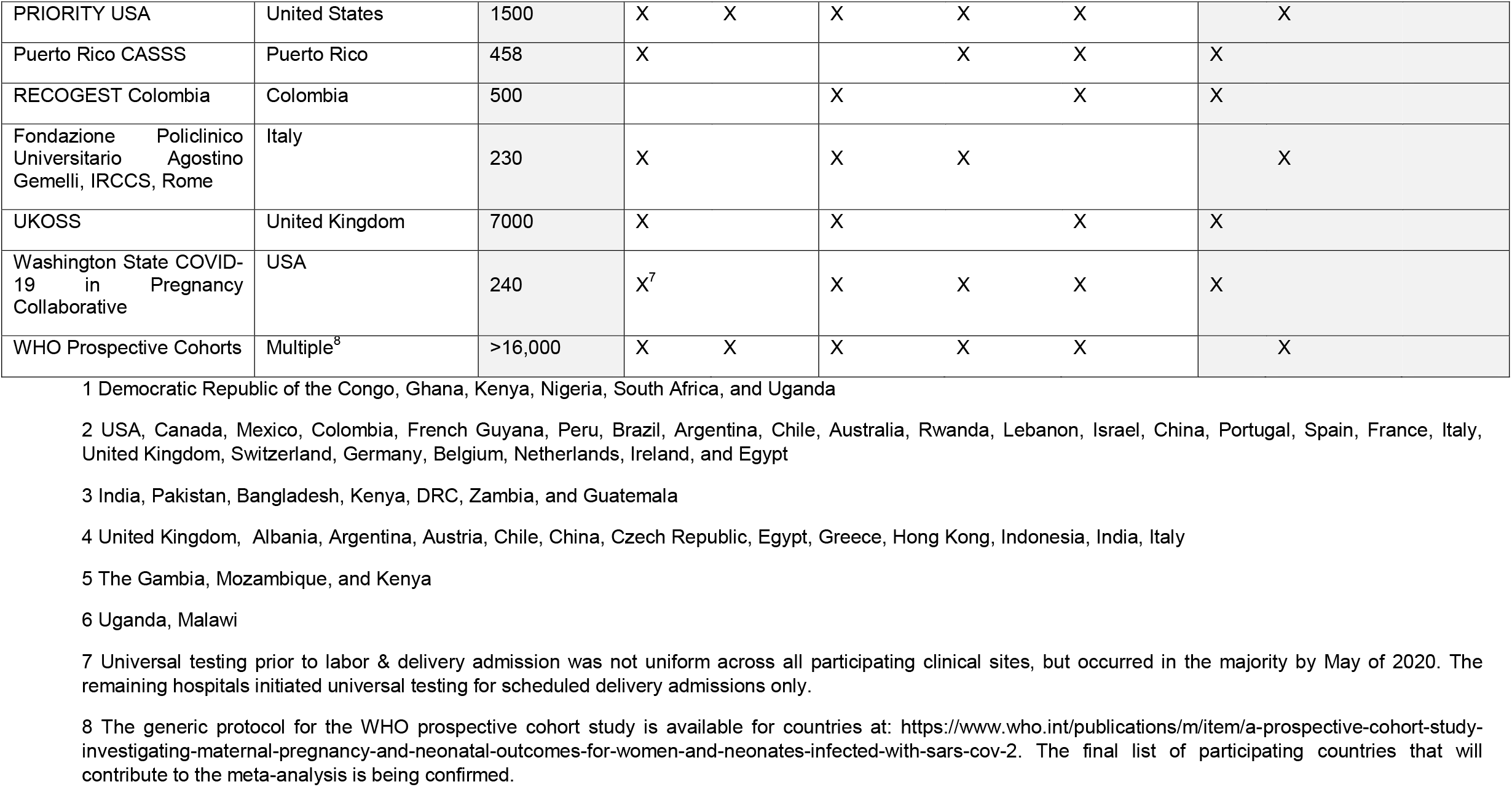
Characteristics of participating studies.

**Fig 1.**
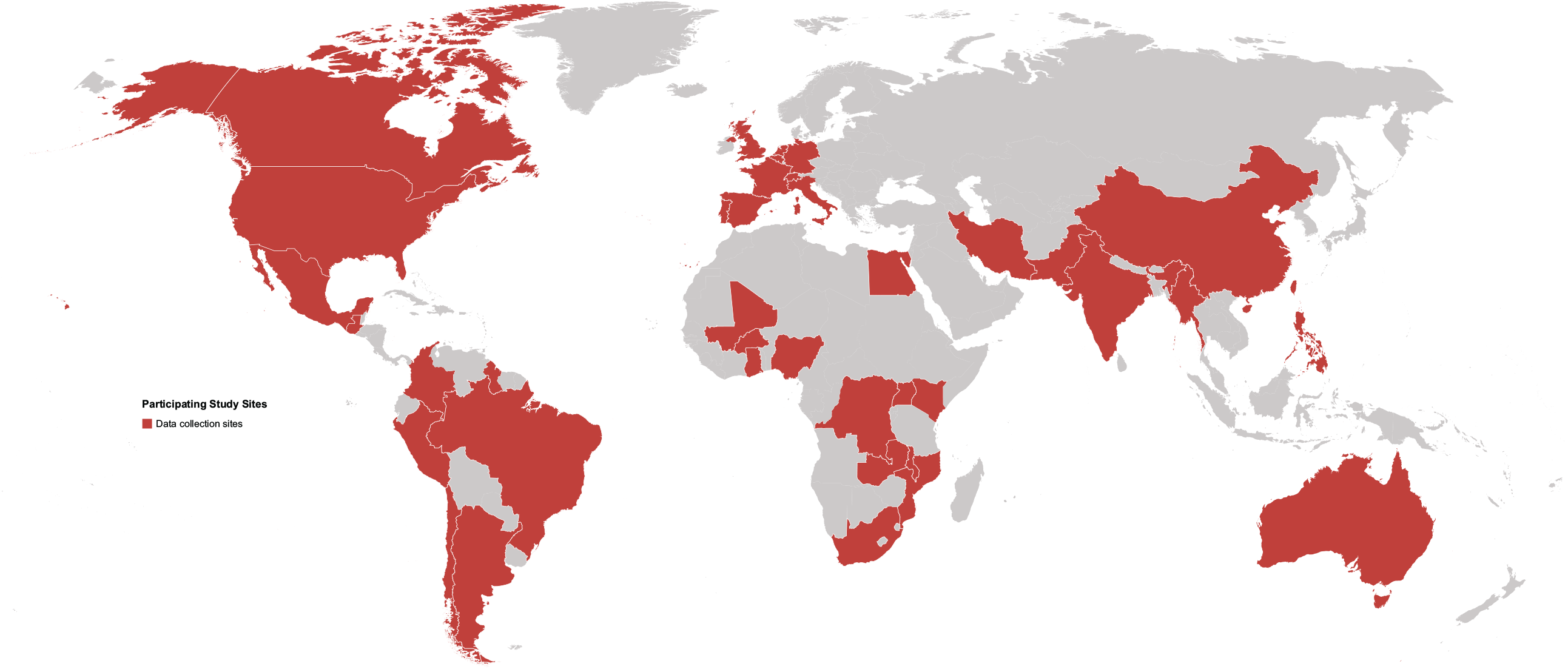
Map of participating study sites.

## Discussion

Our sequential, prospective meta-analysis aims to pool individual patient data from ongoing or completed studies, registries, and surveillance programs that identify SARS-CoV-2 infections in pregnancy and postpartum.

There are several benefits to our approach, which is guided by the principles of flexibility, robustness, collaboration, and inclusion. Early efforts to plan for pooling can reduce research waste (e.g., incompatible data, duplication of efforts) and improve the collective value of data [33]. Through collaboration since April 2020, including participation in WHO-coordinated working groups, we have developed and shared data collection tools and standardized some data collection components while several participating studies were starting their respective research. Ongoing pooling has also highlighted where data elements are missing or sparse across studies, thus providing justification for additional effort from study investigators to retrospectively extract additional data or modify their data collection forms. Further, this collaborative approach of pooling data highlights the possibility of generating robust answers to questions of collective importance at a global scale [34,35]. We are also more likely to elucidate risks related to rare outcomes. Collaboration is especially important for achieving scientific consensus regarding special populations*—*such as pregnant patients and newborns*—*who may not be specifically considered in national surveillance programs and may be actively excluded from other clinical research.

Our approach uses statistical methods for individual patient data meta-analysis, which is generally considered to be the gold standard in pooling data given the many steps used to ensure quality and consistency of the data and analyses. All data will be reviewed with the same data quality assessments, and outliers will be assessed consistently across studies. Analysis definitions will be standardized across studies. This is especially important given that for many outcomes, study definitions vary between countries (e.g. whether a woman should receive critical care, the gestational age cutoff for defining stillbirth). Furthermore, we will avoid duplicate case counting by working with and reanalyzing data from studies that include overlapping health facilities and centers that contribute to two or more studies. This has been a major issue in COVID-19-related meta-analyses because much scientific literature is based on case reports or case series; the same cases sometimes appear in multiple published papers. Traditional meta-analyses based on the published literature cannot resolve these methodological issues.

Importantly, this approach is robust, flexible and collaborative. Each set of investigators can implement their own protocol, offering more flexibility to design a study specific to a given setting. Investigators can also proceed with their own study publications, thus ensuring an opportunity to be explicit about the local context in which the data collection took place and clearly take the credit associated with authorship. Our basic analyses will be done as a two-stage meta-analysis, allowing us flexibility in how data is shared. Studies that are willing and able can share their individual patient data directly with the data coordinating team for central analysis. Alternatively, studies that are unwilling or unable to share individual patient data can appoint their own study-specific data analyst to join the PMA data coordinating team. This approach to a “distributed” individual patient data meta-analysis meets data sharing needs for different groups, and hopefully ensures that anyone can participate. Finally, our regular steering committee meeting allows for joint decision-making about which analyses should be prioritized and when to move forward with publications. We hope to be responsive to emerging, policy-relevant questions, such as the safety and efficacy of vaccination in pregnancy.

## Conclusion

Prospective meta-analyses offer a rigorous way to generate definitive answers to emerging questions. In the current context of the COVID-19 pandemic, there is limited high-quality data that can be pooled in order to inform public health guidance and healthcare strategies, specifically for pregnant women and their newborns. The proposed study will contribute necessary information for evidence-based decision-making related to COVID-19 and maternal and neonatal health.

## Data Availability

This is a protocol paper and there is no related data to share.

## Author Contributions

ES, EO, SH, JMT were involved in the conceptualization and methodology; ES, EO were responsible for data curation; ES, VF, YA were responsible for funding acquisition; ES led investigation and writing the original draft manuscript; EO, SH contributed to the writing – original draft preparation; ES, EO conducted the analysis; EO, RZ contributed to visualization; ES, JMT provided supervision. EO, SH, RZ, KF, LM, and GWG contributed to project administration. All authors contributed to writing – review & editing.

## Supporting information

**S1 File. Data modules and core variables - version June 2, 2020**

**S2 File. Data modules and core variables - version November 8, 2020**

**S3 File. Description of participating studies and key stakeholders**

## Notes

### Competing Interest Statement

Clare Whitehead declares a a relationship with the following entities, Ferring Pharmaceuticals COVID19 Investigational, Grant, NHMRC Fellowship (salary support).
Alice Panchaud declares the following research grants to institution: H2020-Grant (Consortium member of Innovative medicine initiative call 13 topic 9) (ConcePTION), Efficacy and safety studies on Medicines EMA/2017/09/PE/11, Lot 4, WP 2 lead (CONSIGN: Study on impact of COVID-19 infection and medicines in pregnancy), Safety monitoring of COVID-19 vaccines in the EU Reopening of competition no. 20 under a framework contract following procurement procedure EMA/2017/09/PE (Lot 3) 4. Federal Office of Public Health (207000 CHF). (The COVI-Preg registry). Edward Mullins declares a relationship with the following entities National Institute for Health Research (Project grant for PAN COVID study) Deborah Money declares a relationship with the following entities, Canadian Institutes of Health Research (payments to my institution only), Public Health Agency of Canada (payments to my institution only), BC Womens Foundation (payments to my institution only) and is a Member of the COVID-19 Immunity Task Force sponsored by the Canadian government. Torri D. Metz declares a relationship with the following entities, Pfizer (site Principal Investigator for SARS-CoV-2 vaccination in pregnancy study, money paid to institution and member of Medical Advisory Board for SARS-CoV-2 vaccination in pregnancy study, money paid to me), NICHD (subcommittee Chair for the NICHD Maternal-Fetal Medicine Units Network Gestational Research Assessments of COVID-19 (GRAVID) study), and Society for Maternal-Fetal Medicine (board member). Erica Lokken declares a relationship with the following entity, US NIH (paid institution). Karen L. Kotloff declares a relationship with the following entity, Bill and Melinda Gates Foundation. Siran He declares a relationship with the following entity, Bill and Melinda Gates Foundtion (payments made to my institution). Valerie Flaherman declares a relationship with the following entities, Bill and Melinda Gates Foundation (payments to my institution), Yellow Chair Foundation (payments to my institution), Robert Woods Johnson Foundation (payments to my institution), CDC Foundation, California Health Care Foundation (payments to my institution), Tara Health Foundation (payments to my institution), UCSF Womens Health Center of Excellence (payments to my institution) and California Department of Health Care Services (payments made to my institution). Jose Sanin-Blair declares a relationship with the following entity, Ferring Pharmaceuticals which give a grant ($10,000) for the expenses of RECOGEST trial and is a part of the Columbian Federation of Perinatology Yalda Afshar declares a relationship with the following entities, Bill and Melinda Gates Foundation (payments made to my institution), CDC Foundation (payments made to my institution), Robert Woods Johnson Foundation (payments made to my institution), and UCLA Deans Office COVID-19 research (payments made to my institution). Rebecca Cliffton declares a relationship with the following entity, NIH HD36801 (MFMU Network DCC).

### Clinical Trial

PROSPERO ID: 188955

### Funding Statement

Funded by the Bill & Melinda Gates Foundation grant to Emily Smith (INV-022057) at George Washington University and a grant to Emily Smith via a grant from the Bill & Melinda Gates Foundation to Stephanie Gaw (INV-017035) at University of California San Francisco.

### Author Declarations

This is a protocol paper and thus exempt from ethical approval. Ultimately, the meta-analysis study is exempt from human research ethics approval as the study authors will be synthesizing de-identified or aggregate data.

### Summary of Updates

Added feedback from and authorship for the study steering committee.

