## Supplemental Table 1 for "Protocol for a sequential, prospective meta-analysis to describe coronavirus disease 2019 (COVID-19) in the pregnancy and postpartum periods"

### Global Data Harmonization
### Data Modules and Core Questions / Variables for
### Pregnancy & Perinatal COVID-19 Registries or Cohorts

*(Last Updated June 2, 2020)*

*Prepared by Emily R. Smith[1] and Siran He, in collaboration with Yalda Afshar[2] and Valerie Flaherman[3]*

*[1]Department of Global Health, Milken Institute School of Public Health at The George Washington University, Washington, D.C., U.S.A. [2]Department of Obstetrics and Gynecology, University of California, Los Angeles, [3]Department of Pediatrics, University of California, San Francisco*

**Objective:** Our objective is to provide a harmonized set of high priority, core questions / variables for pregnancy and perinatal COVID-19 registry or cohort studies. These are not intended to be a survey or case report form. Each study has or will develop local protocols and data collection forms. However, each study will ideally collect the data outlined here. This will enable future collaborations to answer high priority questions or to pool data where studies investigators are willing and able.

**Harmonization Process:** We developed the draft data modules and questions based on a proposed set of questions from the PRIORITY study. We also reviewed and included questions from the data collection forms developed by the World Health Organization (WHO) and the U.S. Center for Disease Control (CDC). We requested feedback via a survey and by email from the >50 participants of the bi-monthly “Perinatal COVID-19 Global Gathering”. The current data modules reflect feedback and general consensus among survey respondents.

**Which studies should harmonize data?** Any registry or cohort study collecting data regarding pregnant or postpartum people suspected or confirmed to have COVID-19 (as well as their fetus / infant) should consider collecting harmonized data, whether or not they are population-based. The studies may collect data from health records, by directly questioning the participant, or both. Differences in the way participants are sampled/recruited or the way data is collected can be reconciled when specific data analyses are planned.

**Participant Inclusion Criteria:** Participants enrolled in the study should meet the following inclusion criteria:

- Pregnant person, or person who was pregnant within the past 6 weeks *(Note: 42 days/ 6weeks reflects the postpartum period / maternal mortality definition);*
- diagnosed with (or suspected to be infected with) COVID-19;
- provides informed consent.

**What is a Data Module:** We define a "Data Module" as a group of questions (variables) that: 1) are thematically related; 2) are asked at the same time and with the same frequency; 3) AND refer to EITHER mother or baby (not both). These modules are organized for the purpose of prioritizing variables and themes, and do NOT reflect the order in which they should/would appear in actual data collection forms.

**Core Data Modules**

| **Module** | **Research Objective** |
|---|---|
| **Module 1**: Maternal COVID-19 Information *(n=7 questions)* | To evaluate the clinical presentation and natural history of disease for women infected with (or suspected to be infected with) COVID-19 who are pregnant or have been pregnant within the last 6 weeks / 42 days. *(e.g., symptoms, testing, treatment, clinical course)* |
| **Module 2**: Pregnancy Status & Pregnancy-Related Morbidity *(n=16 questions)* | To confirm pregnancy status and to document pregnancy-related basic information among women infected with COVID-19 *(e.g., due date, singleton/multiple pregnancy, etc.)* |
| **Module 3:** Pregnancy Outcomes *(n=6 questions)* | To document the endpoint of the registered pregnancy among women infected with COVID-19 *(e.g., abortion, stillbirth, live birth, etc.)* |
| **Module 4:** Birth Characteristics *(n=14 questions)* | To document various characteristics related to live birth. *(e.g., place of birth, birthweight, gestational age, etc. Information about the infant, if recorded during/right after delivery, will be in this module)* |
| **Module 5:** Infant Morbidity & Mortality *(n=12 questions)* | To evaluate infant outcomes among those born to women who have had COVID-19 (if live birth) *(e.g., COVID-like symptoms up till 12mo, etc.)* |
| **Module 6**: Core socio-demographic information *(n=6 questions)* | To identify high-risk subgroups with increased pregnancy, delivery, and infant adverse outcomes that are potentially associated with COVID-19 |

Time intervals (e.g. gestational age, time from symptom onset to testing) should be calculated directly from dates where possible. **The dates (and ages) recorded in these modules include:**

- Module 1-Q1: Date of onset of COVID-like symptoms
- Module 1-Q2: Date of COVID diagnosis
- Module 2-Q1: Date of study/registry enrollment
- Module 2-Q3: Gestational age upon enrollment
- Module 3-Q1: Date of pregnancy outcome
- Module 3-Q4: Date of maternal death
- Module 4-Q1: Date of birth & time of birth
    - Same as Module 3-Q1 (if live birth)
- Module 4-Q5: Age of infant (in hours) for weighing
- Module 4-Q6: Infant gestational age estimation
    - Can calculate this from Module 3-Q1 (if live birth)
    - Note this question asks about estimation method, which is important in order to address measurement error in analysis stage
- Module 4-Q13: Date of facility discharge after birth (for infant)
- Module 5-Q2: Date of neonatal death
- Module 5-Q6: Date of neonatal COVID testing

#### Module 1: Maternal COVID-19 Information

What: COVID symptoms, testing, treatment, clinical course
When: At regular interval until disease resolution or chart abstraction

| Questionnaire-based data collection | Medical record data extraction |
|---|---|
| Q1. Date of onset of COVID-like symptoms (DD:MM:YY) | Q1. Extract from medical record:<br>● First date of COVID-like symptoms (DD:MM:YY) |
| Q2. Have you been diagnosed with COVID-19?<br>• Yes, confirmed<br>• Not confirmed, under investigation | Q2. Extract from medical record:<br>● COVID-19 diagnosis notes:<br>○ Confirmed<br>○ Patient under investigation (PUI) |
| Q3. If answered “Yes” to Q2:<br>• Date of COVID-19 diagnosis (DD:MM:YY) | Q3. Extract from medical record:<br>• Date of COVID-19 diagnosis, or Date PUI status was documented (DD:MM:YY) |
| Q4. What symptoms did you have that led you to be tested or suspected of Coronavirus/COVID19? (Check all that apply)<br>• Fever<br>• Cough<br>• Shortness of breath<br>• Dizziness or fainting<br>• Body aches<br>• Runny nose<br>• Sore throat<br>• Loss of sense of smell or taste<br>• Sneezing<br>• Fatigue<br>• Nausea<br>• Vomiting<br>• Diarrhea<br>• Headache<br>• Other symptoms (please specify)<br>• None / Asymptomatic | Q4. Extract from medical record:<br>● All symptoms listed in the record that are related to COVID investigation / diagnosis |
| Q5. Do you work in healthcare or provide direct patient care?<br>• Yes<br>• No<br>• Other, please specify | Q5. Extract from medical record:<br>● If occupation data is available,<br>○ Note down “Y” for healthcare/direct patient care,<br>○ “N” for other occupations,<br>○ “NA” for unknown |
| Q6. Have you received any medication for the treatment of COVID-19 (e.g. anti-viral, immunomodulators, convalescent plasma, IL6 mAb, other)<br>• Yes, I have<br>• No, I have not<br>• Maybe / uncertain<br>• Other, please specify | Q6. Extract from medical record:<br>● Has the patient been given any medication for the treatment of COVID-19<br>○ Yes<br>○ No<br>○ Other, please specify |
| Q7. If answered “Yes” to Q6: if possible, document more details about the medication:<br>• Type / Name<br>• Dose<br>• Duration<br>• Indications<br>• Clinical trial registration | Q7. Extract from medical record:<br>• Details about COVID-19 treatment regime, including medications, dose, duration, clinical trial inclusion, etc. |

#### Module 2: Pregnancy Status & Pregnancy-Related Morbidity

What: Pregnancy registration information and non-COVID morbidity among participants
When: At pregnancy registration

| Questionnaire-based data collection | Medical record data extraction |
|---|---|
| Q1. Date of study enrolment (DD:MM:YY) | Q1. Extract from medical record:<br>Date of study enrolment (DD:MM:YY) |
| Q2. Status upon enrolment<br>• Pregnant (confirmed by healthcare provider), not in labor<br>• Pregnant (not confirmed by healthcare provider), not in labor<br>• Pregnant in labor<br>• Postpartum [days] --> if yes, breastfeeding Y/N<br>• Post-abortion, miscarriage | Q2. Extract from medical record:<br>● Pregnancy status:<br>○ Pregnant (confirmed by healthcare provider) not in labor<br>○ Pregnant in labor<br>○ Postpartum [days] --> if yes, breastfeeding Y/N<br>○ Post-abortion, miscarriage |
| Q3. If answered "postpartum" in Q2: are you breastfeeding?<br>• Yes<br>• No | Q3. Extract from medical record:<br>• Breastfeeding status for postpartum women |
| Q4. Gestational age at enrollment. Do you know your due date?<br>• Yes → document due date (or how far along you are in pregnancy) (DD:MM:YY)<br>• No<br>• Maybe / uncertain | Q4. Extract from medical record:<br>● Gestation age at enrollment |
| Q5. If answered "Yes" to Q4:<br>• What is the method of due date assessment? | Q5. Extract from medical record:<br>• Method of gestational age assessment |
| Q6. Number of fetuses:<br>• Singleton<br>• Twins<br>• Other, specify | Q6. Extract from medical record:<br>● Number of fetuses |
| Q7. (Pre-pregnancy) BMI (can document BMI or weight and height):<br>• Weight (Kg)<br>• Height (m) | Q7. Extract from medical record:<br>● Pre-pregnancy weight and height (if available, be sure to note down units) |
| Q8. (At the time of registration) BMI (can document BMI or weight and height):<br>• Weight (Kg)<br>• Height (m) | Q8. Extract from medical record:<br>• At the time of clinical visit, weight and height (if available, note down units) |
| Q9. Gravidity: Is this your first pregnancy?<br>• Yes<br>• No | Q9. Extract from medical record:<br>● Gravidity information |
| Q10. If answered "No" to Q9:<br>• How many times have you been pregnant? | Q10. NA |
| Q11. Parity: Is this your first child birth?<br>• Yes<br>• No | Q11. Extract from medical record:<br>● Parity information |
| Q12. If answered "Not" to Q11:<br>• How many live births have you had previously? | Q12. NA |

| | |
|---|---|
| Q13. Has a doctor or other healthcare provider told you that you have any of the following conditions before you were pregnant? (check all that apply) *(local sites should describe conditions in a way that women will understand and self-report)*<br>• Asthma<br>• Obesity<br>• Sleep apnea<br>• Anemia (Hb < 11g/dL per WHO)<br>• Chronic high blood pressure (hypertension)<br>• Thyroid disease<br>• Immune suppression (due to underlying disease or meds)<br>• Neurological disease<br>• Chronic lung disease (excluding asthma)<br>• Autoimmune disease<br>• Cardiovascular disease (excluding hypertension)<br>• Other, please note | Q13. Extract from medical record:<br>• List all pregnancy-related conditions documented in the record |
| Q14. Has a doctor or other healthcare provider told you that you have any of the following conditions during pregnancy? (check all that apply) *(local sites should describe conditions in a way that women will understand and self-report)*<br>• Hypertensive disease of pregnancy (including preeclampsia/eclampsia)<br>• Hyperemesis<br>• Intrauterine growth restriction<br>• Abnormal placentation (placental previa/accreta/percreta)<br>• Placental abruption<br>• Bacterial infection prior to hospital visit<br>• Preterm contractions (not in labor)<br>• Preterm labor<br>• Preterm rupture of membranes<br>• Haemorrhage<br>• If haemorrhage, which type: antepartum/intrapartum; Postpartum; Abortion-related<br>• Embolic disease<br>• Anesthetic complications | Q14. Extract from medical record:<br>• List all pregnancy-related conditions documented in the record |
| Q15. If any sample was collected for research, what was/were the sample(s)?<br>• Amniotic fluid<br>• Placenta<br>• Cord blood<br>• Vaginal swab<br>• Feces/rectal swab<br>• Pregnancy tissue in the case of fetal demise/induced abortion<br>• Breastmilk<br>• Maternal blood<br>• Not applicable (no sample collected) | Q15. Extract from medical record:<br>• Document collection of any biological sample |
| Q16. If selected any sample(s) in Q15:<br>• If possible, please document for each type of sample collected:<br> ○ Tests done with the samples<br> ○ Results of the tests | Q16. Extract from medical record:<br>• Separately for each sample documented:<br> ○ Tests done<br> ○ Results of the tests |

#### Module 3: Pregnancy Outcomes

What: Information about how the pregnancy ended; maternal mortality will be recorded in this module
When: Once, after pregnancy outcome is known

| **Questionnaire-based data collection** | **Medical record data extraction** |
|---|---|
| Q1. Date of pregnancy outcome (DD:MM:YY)<br>*[Allows calculation of gestational age (in weeks) when pregnancy ended]* | Q1. Extract from medical record:<br>● Date of pregnancy outcome (DD:MM:YY) |
| Q2. Please indicate the end point of this pregnancy:<br>• Live birth<br>• Stillbirth<br>• SAB (spontaneous abortion) → expectant, medical management, D&C/E (3 choices)<br>• TAB (therapeutic abortion) → expectant, medical management, D&C/E (3 choices) | Q2. Extract from medical record:<br>● The end point of this pregnancy (note down based on the format in the record, such as live birth, stillbirth, etc.) |
| Q3. Maternal death?<br>• Yes<br>• No<br>• Unknown | Q3. Extract from medical record:<br>● If maternal death occurred (yes/no/unknown) |
| Q4. If answered "Yes" for Q3:<br>• Date of maternal death? (DD:MM:YY) | Q4. Extract from medical record:<br>• If maternal death, date of death (DD:MM:YY) |
| Q5. If answered "Yes" for Q3:<br>• Gestational age in weeks at death? | Q5. Extract from medical record:<br>• If maternal death, gestational age in weeks at death |
| Q6. If answered "Yes" for Q3:<br>• Cause of death<br>o COVID-19<br>o Obstetric hemorrhage<br>o Hypertensive disorder (including preeclampsia and eclampsia)<br>o Pregnancy-related infection<br>o Abortion/ectopic pregnancy<br>o Other direct cause (obstetric complications)<br>o Indirect cause (pre-existing medical condition exacerbated by pregnancy)<br>o Coincidental cause (e.g. motor vehicle cause, accidental injury, assault)<br>o Unknown | Q6. Extract from medical record:<br>• If maternal death, cause of maternal death (COVID or other causes, document as appeared in medical record) |

#### Module 4: Birth Characteristics

What: Information related to labor and delivery/birth and other data collected on the day of birth
When: To be asked on or soon after the day of birth, at enrollment if enrollment occurs postpartum, or directly extracted from chart

| **Questionnaire-based data collection** | **Medical record data extraction** |
|---|---|
| Q1. Date (and time, if available) of birth (DD:MM:YY) (HH:MM) | Q1. Extract from medical record:<br>• Date of birth (and time of birth, if available) (DD:MM:YY) (HH:MM) |
| Q2. Where was the infant born? *<br>• Home<br>• Birthing facility/hospital<br>• Other location, please specify<br><br>** If your registry collects only hospital-based births, please record "facility/hospital" for all records* | Q2. Extract from medical record:<br>• Location of birth:<br>○ Hospital (indicate which hospital)<br>○ Other (specify location, if delivery did not occur at this facility)<br>○ "NA" if unknown |
| Q3. Mode of delivery?<br>• Spontaneous vaginal<br>• Operative vaginal - forceps or vacuum<br>• Cesarean section (scheduled)<br>• Cesarean section (intrapartum)<br>• Unknown | Q3. Extract from medical record:<br>• Mode of delivery exactly as documented in the record |
| Q4. What was the infant's weight at birth (grams)? | Q4. Extract from medical record:<br>• Infant weight at birth (grams) |
| Q5. Has the infant's gestational age been estimated?<br>• Yes<br>• No | Q5. NA |
| Q6. If answered "Yes" to Q6:<br>• What was the estimated gestational age (weeks) | Q6. Extract from medical record:<br>• Gestational age of infant |
| Q7. If answered "Yes to Q6:<br>• What method was used to determine gestational age?<br>○ Ultrasound<br>○ Estimated date of delivery based on last menstrual period (EDD)<br>○ Ballard/Dubowitz;<br>○ Other, please specify | Q7. Extract from medical record:<br>• Estimation method exactly as documented in the record |
| Q8. On the day of birth, did your infant have any difficulty breathing?<br>• Yes<br>• No<br>• Maybe / Uncertain | Q8. Extract from medical record:<br>• On the day of birth, did the infant have any difficulty breathing, as documented in the record |
| Q9. At the time of birth, was your infant admitted to the neonatal intensive care unit or the special care unit?<br>• Yes<br>• No | Q9. Extract from medical record:<br>• At the time of birth, was the infant admitted to the neonatal intensive care unit/special care unit (Y/N/NA) |
| Q10. At the time of birth, did your infant receive oxygen?<br>• Yes<br>• No | Q10. Extract from medical record:<br>• At the time of birth, did the infant receive oxygen? (Y/N/NA) |

| | |
|---|---|
| Q11. What type of resuscitation was provided at delivery?<br>• None<br>• Warm/dry/stimulation<br>• Blow by oxygen<br>• Continuous positive airway pressure<br>• Positive pressure ventilation<br>• Intubation<br>• Chest compressions<br>• Surfactant<br>• Unknown | Q11. Extract from medical record:<br>• Resuscitation? (Y/N/NA)<br>• If yes → type of resuscitation, as documented in the record |
| Q12. Did the infant breastfeed or receive any breast milk on the day of birth?<br>• Yes<br>• No<br>• Maybe / Uncertain | Q12. Extract from medical record:<br>• Breastfeeding or breast milk on the day of birth (Y/N/NA) |
| Q13. Date of infant discharge from labor and delivery event (DD:MM:YY) | Q13. Extract from medical record:<br>• Date of infant discharge (DD:MM:YY) |
| Q14. Newborn outcome at discharge:<br>• Expired<br>• Home<br>• Transfer to another acute care facility<br>• Transfer to a chronic care facility | Q14. Extract from medical record:<br>• Newborn outcomes at discharge as documented in the record |

#### Module 5: Infant Mortality and Morbidity

What: Information related to infant health, particularly related to COVID-19
When: At regular interval until up to 12 months after birth (or until chart extraction)

| Questionnaire-based data collection | Medical record data extraction |
|---|---|
| Q1. Neonatal/infant death?<br>• Yes<br>• No | Q1. Extract from medical record:<br>• Neonatal/infant death? (Y/N/NA) |
| Q2. If answered “Yes” to Q1:<br>• Date-time of neonatal/infant death? (DD:MM:YY) (HH:MM) | Q2. Extract from medical record:<br>• If neonatal/infant death, date-time of death (DD:MM:YY) (HH:MM) |
| Q3. If answered “Yes” to Q1:<br>• Cause of death?<br>o COVID-19<br>o Preterm/low birth weight<br>o Birth asphyxia<br>o Infection<br>o Birth trauma<br>o Congenital/birth defects<br>o Unknown<br>o Other, please specify | Q3. Extract from medical record:<br>• If neonatal/infant death, cause of death (COVID or other causes, document as appeared in medical record) |
| Q4. Was the infant documented by clinicians to have any of the following symptoms of COVID-19?<br>- Fever (T>37.5)<br>- Respiratory distress<br>- Cough<br>- Nasal congestion/runny nose<br>- Vomiting<br>- Diarrhea<br>- Lethargy<br>- Rapid heart rate (>160 bpm) (record bpm if available) | Q4. Extract from medical record:<br>• Was the infant documented by clinicians to have any of the symptoms of COVID-19? (document exactly as in the record) |
| Q5. Was the baby tested for COVID-19?<br>• Yes<br>• No<br>• Maybe / Uncertain | Q5. Extract from medical record:<br>• Any COVID-19 test for the infant? (Y/N) |
| Q6. If answered “Yes” to Q5:<br>• When was the baby tested?<br>o Immediately after birth<br>o Other, please specify date (DD:MM:YY) (HH:MM) | Q6. Extract from medical record:<br>• Testing date and time (DD:MM:YY) (HH:MM) |
| Q7. If answered “Yes” to Q5:<br>• What tests were conducted?<br>o Viral PCR test<br>o Chest image or X-ray<br>o Other, please specify | Q7. Extract from medical record:<br>• Which tests were conducted |
| Q8. If answered “Yes” to Q5:<br>• What was the test result?<br>o Confirmed COVID-19<br>o Not COVID-19<br>o Other, specify | Q8. Extract from medical record:<br>• Test result – exactly as documented |
| Q9. Has the baby received antiviral medications? | Q9. Extract from medical record: |

| | |
|---|---|
| → If so, which antiviral medication was administered? | • Did the infant receive antiviral medication? (Y/N/NA) |
| Q10. If answered “Yes” to Q9:<br>• Which antiviral medication was administered? If possible, document:<br> ○ Name<br> ○ Dosage<br> ○ Duration | Q10. Extract from medical record:<br>• If antiviral medication was used, note down exactly as in the record:<br> ○ Which medication<br> ○ Dosage<br> ○ Duration |
| Q11. Has your infant breastfed or received any breast milk in the last 24 hours?<br>• Yes<br>• No<br>• Other, please specify | Q11. Extract from medical record:<br>• Has the infant been breastfed or received any breast milk in the past 24 hours (Y/N/NA)?<br>• If information is available for longer periods, note it down as well |
| Q12. Any congenital anomalies?<br>• Neural tube defects<br>• Microcephaly<br>• Congenital malformations of ear<br>• Congenital heart defects<br>• Orofacial clefts<br>• Congenital malformations of digestive system<br>• Congenital malformations of genital organs<br>• Abdominal wall defects<br>• Chromosomal abnormalities<br>• Reduction defects of upper and lower limbs<br>• Talipes equinovarus/clubfoot | Q12. Extract from medical record:<br>• Any congenital anomalies in the record? (Note it down exactly as in the record) |

#### Module 6: Core Sociodemographic Information

What: Socio-demographic information about the study participant
When: Once at the beginning of the study (maternal information); Once after birth (for infant information)

| Questionnaire-based data collection | Medical record data extraction |
|---|---|
| Q1. Country \| Site ID | Q1. Indicate the site / facility |
| Q2. Age of the mother (years) | Q2. Extract from medical record:<br>● Mother's age (years), or<br>● Mother's date of birth (DD:MM:YY) |
| Q3. Sex of the child (if applicable) | Q3. Extract from medical record:<br>● Sex of the child |
| Q4. Race/Ethnicity of the mother *<br><br>** Each site to define appropriate categories for their context* | Q4. Extract from medical record:<br>● Mother's race/ethnicity (if available) |
| Q5. Race/Ethnicity of the child (if applicable) *<br><br>** Each site to define appropriate categories for their context* | Q5. Extract from medical record:<br>● Child's race/ethnicity (if available) |
| Q6. Years/level of education completed by the mother (check highest level completed)<br>• No formal education<br>• Some primary education<br>• Primary education completed<br>• Secondary education completed<br>• University/College completed<br>• Graduate education /Terminal degree completed | Q6. Extract from medical record:<br>● Mother's education level (if available) |
