## Supplemental Table 2 for "Protocol for a sequential, prospective meta-analysis to describe coronavirus disease 2019 (COVID-19) in the pregnancy and postpartum periods"

### Global Data Harmonization
### Data Modules and Core Questions / Variables for Pregnancy & Perinatal COVID-19 Registries or Cohorts

*(Last Updated November 8, 2020)*

*Prepared by Emily R. Smith[1] and Siran He[1], in collaboration with Yalda Afshar[2] and Valerie Flaherman[3]*

*Updated by Emily R. Smith[1], Siran He[1], Erin Oakley[1], Mollie Wood[4]*

*[1]Department of Global Health, Milken Institute School of Public Health at The George Washington University, Washington, D.C., U.S.A. [2]Department of Obstetrics and Gynecology, University of California, Los Angeles, [3]Department of Pediatrics, University of California, San Francisco, [4]Cincinnati College of Medicine, Cincinnati*

**Updates**: Updates to the data modules in August and September 2020 reflect efforts to harmonize these modules with updated World Health Organization (WHO) case definitions and ongoing multi-site study protocols developed by WHO. Additions are highlight in yellow. Questions that have been removed are crossed out.

#### Core Data Modules

| Module | Research Objective |
|---|---|
| **Module 1**: Maternal COVID-19 Information<br>*(n=14 questions)* | To evaluate the clinical presentation and natural history of disease for women infected with (or suspected to be infected with) COVID-19 who are pregnant or have been pregnant within the last 6 weeks / 42 days. *(e.g., symptoms, testing, treatment, clinical course)* |
| **Module 2**: Pregnancy Status & Pregnancy-Related Morbidity<br>*(n=14 questions)* | To confirm pregnancy status and to document pregnancy-related basic information among women infected with COVID-19<br>*(e.g., due date, singleton/multiple pregnancy, etc.)* |
| **Module 3:** Pregnancy Outcomes<br>*(n=5 questions)* | To document the endpoint of the registered pregnancy among women infected with COVID-19<br>*(e.g., abortion, stillbirth, live birth, etc.)* |
| **Module 4:** Birth Characteristics<br>*(n=15 questions)* | To document various characteristics related to live birth.<br>*(e.g., place of birth, birthweight, gestational age, etc. Information about the infant, if recorded during/right after delivery, will be in this module)* |
| **Module 5:** Infant Morbidity & Mortality<br>*(n=8 questions)* | To evaluate infant outcomes among those born to women who have had COVID-19 (if live birth)<br>*(e.g., COVID-like symptoms up till 12mo, etc.)* |
| **Module 6**: Core socio-demographic information<br>*(n=6 questions)* | To identify high-risk subgroups with increased pregnancy, delivery, and infant adverse outcomes that are potentially associated with COVID-19 |
| **Module 7:** Biospecimens and Diagnostic Testing<br>*(n=unlimited responses based on collected samples)* | To clearly document all biospecimens collected related to COVID-19 in a pregnancy and delivery and diagnostic testing for COVID-19 completed for the mother and infant, with a focus on identifying any instances of vertical transmission of COVID-19. |

Time intervals (e.g. gestational age, time from symptom onset to testing) should be calculated directly from dates where possible. **The dates (and ages) recorded in these modules include:**

- Module 1-Q1: Date of onset of COVID-like symptoms
- Module 1-Q3: Date of COVID diagnosis
- Module 1-Q8: Date of hospital admission
- Module 1-Q13: Date of ICU admission / critical care receipt
- Module 2-Q1: Date of study/registry enrollment
- Module 2-Q4: Expected date of delivery
- Module 3-Q1: Date of pregnancy outcome
- Module 3-Q4: Date of maternal death
- Module 4-Q1: Date of birth & time of birth
    - Same as Module 3-Q1 (if live birth)
- Module 4-Q6: Infant gestational age estimation
    - Note this question asks about estimation method, which is important in order to address measurement error in analysis stage
- Module 4-Q14: Date of facility discharge after birth (for infant)
- Module 5-Q2: Date of neonatal death
- Module 5-Q4: Date of neonatal COVID-19 signs/symptoms
- Module 7-Q2a: Collection date of maternal biospecimen(s)
- Module 7-Q4a: Collection date of biospecimen(s) at delivery
- Module 7-Q6a: Collection date of neonatal/infant biospecimen(s)

#### Module 1: Maternal COVID-19 Information

What: COVID symptoms, testing, treatment, clinical course
When: At regular interval until disease resolution or chart abstraction

| Questionnaire-based data collection | Medical record data extraction |
|---|---|
| Q1. Date of onset of COVID-like symptoms (DD:MM:YY) | Q1. Extract from medical record:<br>● First date of COVID-like symptoms (DD:MM:YY) |
| Q2. Have you been diagnosed with COVID-19?<br>● Yes, laboratory confirmed COVID-19<br>● Yes, probable COVID-19 case (see 2a)<br>● Suspected COVID-19 case (see 2b)<br>● No, COVID-19 negative | Q2. Extract from medical record:<br>● COVID-19 diagnosis notes:<br>○ Laboratory Confirmed<br>○ Probable COVID-19 case (see 2a)<br>○ Suspected COVID-19 case (see 2b)<br>○ No, COVID-19 negative |
| Q2a. If diagnosed as a probable COVID-19 case, by which criteria where you diagnosed?<br>• A patient who meets clinical criteria above AND is a contact of a probable or confirmed case, or epidemiologically linked to a cluster with at least one confirmed case. | Q2a. If diagnosed as a probable COVID-19 case, by which criteria was diagnosis made?<br>• A patient who meets clinical criteria above AND is a contact of a probable or confirmed case, or epidemiologically linked to a cluster with at least one confirmed case. |

| | |
|---|---|
| • A suspect case with chest imaging showing findings suggestive of COVID-19 disease<br>• A person with recent onset of anosmia (loss of smell) or ageusia (loss of taste) in the absence of any other identified cause.<br>• Death, not otherwise explained, in an adult with respiratory distress preceding death AND was a contact of a probable or confirmed case or epidemiologically linked to a cluster with at least one confirmed case. | • A suspect case with chest imaging showing findings suggestive of COVID-19 disease<br>• A person with recent onset of anosmia (loss of smell) or ageusia (loss of taste) in the absence of any other identified cause.<br>Death, not otherwise explained, in an adult with respiratory distress preceding death AND was a contact of a probable or confirmed case or epidemiologically linked to a cluster with at least one confirmed case. |
| Q2b. If diagnosed as a probable COVID-19 case, by which criteria where you diagnosed?<br>• A person who meets the clinical AND epidemiological criteria by WHO<br>• A patient with severe acute respiratory illness | Q2b. If diagnosed as a probable COVID-19 case, by which criteria was diagnosis made?<br>• A person who meets the clinical AND epidemiological criteria by WHO<br>A patient with severe acute respiratory illness |
| Q3. If answered "Yes" to Q2:<br>• Date of COVID-19 diagnosis (DD:MM:YY) | Q3. Extract from medical record:<br>• Date of COVID-19 diagnosis, or Date PUI status was documented (DD:MM:YY) |
| Q4. What symptoms did you have that led you to be tested or suspected of Coronavirus/COVID19? (Check all that apply)<br>• Fever<br>• Cough<br>• Shortness of breath<br>• Dizziness or fainting<br>• Body aches<br>• Runny nose<br>• Sore throat<br>• Loss of sense of smell<br>• Loss of sense of taste<br>• Sneezing<br>• Fatigue<br>• Nausea<br>• Vomiting<br>• Diarrhea<br>• Headache<br>• Other symptoms (please specify)<br>• None / Asymptomatic | Q4. Extract from medical record:<br>• All symptoms listed in the record that are related to COVID investigation / diagnosis |
| Q5. Do you work in healthcare or provide direct patient care?<br>• Yes<br>• No<br>• Other, please specify | Q5. Extract from medical record:<br>• If occupation data is available,<br>○ Note down "Y" for healthcare/direct patient care,<br>○ "N" for other occupations,<br>○ "NA" for unknown |
| Q6. Have you received any medication for the treatment of COVID-19 (e.g. anti-viral, immunomodulators, convalescent plasma, IL6 mAb, other)<br>• Yes, I have<br>• No, I have not<br>• Maybe / uncertain<br>• Other, please specify | Q6. Extract from medical record:<br>• Has the patient been given any medication for the treatment of COVID-19<br>○ Yes<br>○ No<br>○ Other, please specify |

| | |
|---|---|
| Q7. If answered "Yes" to Q6: if possible, document more details about the medication:<br>● Type / Name<br>● Dose<br>● Duration<br>● Indications<br>● Clinical trial registration | Q7. Extract from medical record:<br>● Details about COVID-19 treatment regime, including medications, dose, duration, clinical trial inclusion, etc. |
| Q8. Were you ever admitted to the hospital for COVID-19<br>• Yes<br>• No<br>• Unknown | Q8. Extract from medical record: Ever admitted to the hospital for COVID-19<br>• Yes<br>• No<br>Unknown |
| Q9. If answer "Yes" to Q8": What was the date of hospital admission? (DD:MM:YY) | Q9. Extract from medical record: What was the date of hospital admission? (DD:MM:YY) |
| Q10. If answer "Yes" to Q8": What was the date of hospital discharge? (DD:MM:YY) (in the case of death, enter date of death). | Q10. If answer "Yes" to Q8": Extract from medical record: What was the date of hospital discharge? (DD:MM:YY) (in the case of death, enter date of death). |
| Q11. If answer "Yes" to Q8": what was the respiratory status of the patient while hospitalized?<br>• Self ventilating in room air<br>• Self ventilating with oxygen support<br>• Non-invasive respiratory support (CPAP, NIV)<br>• Mechanically ventilated, intubation<br>• Unknown | Q11. If answer "Yes" to Q8": Extract from medical record: what was the respiratory status of the patient while hospitalized?<br>• Self ventilating in room air<br>• Self ventilating with oxygen support<br>• Non-invasive respiratory support (CPAP, NIV)<br>• Mechanically ventilated, intubation<br>• Unknown |
| Q12. If answer "Yes" to Q8": Was the mother admitted to an intensive care unit (ICU) or administered critical care for COVID-19? | Q12. If answer "Yes" to Q8": Extract from medical record: Was the patient admitted to an intensive care unit (ICU) or administered critical care for COVID-19? |
| Q13. If answer "Yes" to Q12": What was the date of ICU admission / critical care? (DD:MM:YY) | Q13. If answer "Yes" to Q12": Extract from medical record: What was the date of ICU admission / critical care? (DD:MM:YY) |
| Q14. If answer "Yes" to Q8": What was the date of ICU discharge / end of critical care? (DD:MM:YY) (in the case of death, enter date of death). | Q14. If answer "Yes" to Q8": Extract from medical record: What was the date of ICU discharge / end of critical care? (DD:MM:YY) (in the case of death, enter date of death). |

| | |
|---|---|
| ● Unknown | |
| Q12. Did the infant breastfeed or receive any breast milk on the day of birth?<br>● Yes<br>● No<br>● Maybe / Uncertain | Q12. Extract from medical record:<br>● Breastfeeding or breast milk on the day of birth (Y/N/NA) |
| Q13: Was the newborn isolated away from mother in another area in hospital (postnatal ward, special care nursery, NICU or special ward)? | Q13: Extract from medical record:<br>● Was the newborn isolated away from mother in another area in hospital (postnatal ward, special care nursery, NICU or special ward)? |
| Q14. Date of infant discharge from labor and delivery event (DD:MM:YY) | Q14. Extract from medical record:<br>● Date of infant discharge (DD:MM:YY) |
| Q15. Newborn outcome at discharge:<br>● Expired<br>● Home<br>● Transfer to another acute care facility due to clinical needs<br>● Transfer to a chronic care facility | Q15. Extract from medical record:<br>● Newborn outcomes at discharge as documented in the record |

**Module 5: Infant Mortality and Morbidity**

What: Information related to infant health, particularly related to COVID-19
When: At regular interval until up to 12 months after birth (or until chart extraction)

| **Questionnaire-based data collection** | **Medical record data extraction** |
|---|---|
| Q1. Neonatal/infant death?<br>● Yes<br>● No | Q1. Extract from medical record:<br>● Neonatal/infant death? (Y/N/NA) |
| Q2. If answered “Yes” to Q1:<br>● Date-time of neonatal/infant death? (DD:MM:YY) (HH:MM) | Q2. Extract from medical record:<br>● If neonatal/infant death, date-time of death (DD:MM:YY) (HH:MM) |
| Q3. If answered “Yes” to Q1:<br>● Cause of death?<br>○ COVID-19<br>○ Preterm/low birth weight<br>○ Birth asphyxia<br>○ Infection<br>○ Birth trauma<br>○ Congenital/birth defects<br>○ Unknown<br>○ Other, please specify | Q3. Extract from medical record:<br>● If neonatal/infant death, cause of death (COVID or other causes, document as appeared in medical record) |
| Q4. Was the infant documented by clinicians to have any of the following symptoms of COVID-19?<br>- Fever (T>37.5)<br>- Respiratory distress | Q4. Extract from medical record:<br>● Was the infant documented by clinicians to have any of the symptoms of COVID-19? (document exactly as in the record) |

| | |
|---|---|
| - Cough<br>- Nasal congestion/runny nose<br>- Vomiting<br>- Diarrhea<br>- Lethargy<br>- Rapid heart rate (>160 bpm) (record bpm if available)<br>- Seizure<br>- Paralysis<br>- Hypotonia (floppiness)<br>- Hypertonia or Stiffness or spasticity of limbs<br>- Other neurological signs<br>- Rash<br>- Oedema<br>- Eye redness/conjunctivitis<br>- Other condition<br><br>*If yes, date of onset?* | |
| Q5. Was the baby tested for COVID-19?<br>● Yes<br>● No<br>● Maybe / Uncertain | Q5. Extract from medical record:<br>● Any COVID-19 test for the infant? (Y/N)<br><br>*If answered "Yes" go to Module 7 to record details.* |
| Q6. If answered "Yes" to Q5:<br>● What tests were conducted?<br>○ Biospecimen: viral, antibody, or antigen test (record details in Module 7)<br>○ Chest image or X-ray<br>○ Lung ultrasound<br>○ Echcardiogram<br>○ Cerebral ultrasound<br>○ Abdominal ultrasound<br>○ Other, please specify<br><br>*If answered "Yes" to "Biospecimen" - go to Module 7 to record details.* | Q6. Extract from medical record:<br>● Which tests were conducted |
| Q7. Has your infant breastfed or received any breast milk in the last 24 hours?<br>● Yes<br>● No<br>● Unknown | Q7. Extract from medical record:<br>● Has the infant been breastfed or received any breast milk in the past 24 hours (Y/N/NA)?<br>● If information is available for longer periods, note it down as well |
| Q8. Any congenital anomalies?<br>● Neural tube defects<br>● Microcephaly<br>● Congenital malformations of ear<br>● Congenital heart defects<br>● Orofacial clefts<br>● Congenital malformations of digestive system<br>● Congenital malformations of genital organs<br>● Abdominal wall defects<br>● Chromosomal abnormalities<br>● Reduction defects of upper and lower limbs | Q8. Extract from medical record:<br>● Any congenital anomalies in the record? (Note it down exactly as in the record) |

| |
|---|
| • Talipes equinovarus/clubfoot<br>• Other, please specify |

#### Module 6: Core Sociodemographic Information

What: Socio-demographic information about the study participant
When: Once at the beginning of the study (maternal information); Once after birth (for infant information)

| Questionnaire-based data collection | Medical record data extraction |
|---|---|
| Q1. Country \| Site ID | Q1. Indicate the site / facility |
| Q2. Age of the mother (years) | Q2. Extract from medical record:<br>• Mother's age (years), or<br>• Mother's date of birth (DD:MM:YY) |
| Q3. Sex of the child (if applicable)<br>• Male<br>• Female<br>• Ambiguous<br>• Unknown | Q3. Extract from medical record:<br>• Sex of the child |
| Q4. Race/Ethnicity of the mother *<br><br>** Each site to define appropriate categories for their context* | Q4. Extract from medical record:<br>• Mother's race/ethnicity (if available) |
| Q5. Race/Ethnicity of the child (if applicable) *<br><br>** Each site to define appropriate categories for their context* | Q5. Extract from medical record:<br>• Child's race/ethnicity (if available) |
| Q6. Years/level of education completed by the mother (check highest level completed)<br>• No formal education<br>• Some primary education<br>• Primary education completed<br>• Secondary education completed<br>• University/College completed<br>• Graduate education /Terminal degree completed | Q6. Extract from medical record:<br>• Mother's education level (if available) |

#### Module 7: Biospecimens and Diagnostic Testing

What: Biological specimens collected from mother and infant during pregnancy, at delivery, and postpartum, with a focus on indicators of vertical transmission of COVID-19.
When: Maternal biospecimens collected during the pregnancy, at the time of delivery, or after delivery, as well as biospecimens collected during delivery and from the infant at birth and during the first 6 weeks of life.

| **Questionnaire-based data collection** | **Medical record data extraction** |
|---|---|
| Q1. Were any biospecimens collected for COVID-19 diagnostic testing from the mother during pregnancy, at the time of delivery, or in the postpartum period? | Q1. Extract from medical record:<br>• Any biospecimen/diagnostic testing for the mother? (Y/N) |
| Q2. If answered "Yes" to Q1: → Proceed to maternal biospecimen table below | See Q2 Maternal Biospecimen Table below |
| Q3. Were any biospecimens collected for COVID-19 diagnostic testing at the time of the delivery? | Q3. Extract from medical record:<br>• Any biospecimen/diagnostic testing at delivery? (Y/N) |
| Q4. If answered "Yes" to Q3: → Proceed to delivery-related biospecimen table below | See Q3 Delivery-Related Biospecimen Table below |
| Q5. Were any biospecimens collected for COVID-19 diagnostic testing from the infant? | Q5. Extract from medical record:<br>Any biospecimen/diagnostic testing for the infant? (Y/N) |
| Q6. If answered "Yes" to Q5: → Proceed to infant biospecimen table below | See Q3 Infant Biospecimen Table below |

Q2. Maternal Biospecimen Table

| Sample Number (as needed) | 2a. Date of Biospecimen Collection | 2b. Type of Biospecimen | 2c. Type of Testing Conducted | 2d. Qualitative Results | 2e. Quantitative Result (e.g., viral load) | 2f. Specify units for Quantitative Results |
|---|---|---|---|---|---|---|
| SM1. | DD:MM:YY | • Nasopharyngeal swab<br>• Vaginal swab<br>• Feces/rectal swab<br>• Maternal blood<br>• Breast milk<br>• Pregnancy tissue (in the case of fetal demise/induced abortion)<br>• Other, specify | • Viral PCR<br>• IgM<br>• IgG<br>• Other, specify | • Positive<br>• Negative<br>• Other, specify | ___ | ________<br>________ |
| SM2. | | | | | | |
| SM3. | | | | | | |
| SM4. | | | | | | |

Q4. Delivery-Related Biospecimen Table

| Sample Number (as needed) | 4a. Date of Biospecimen Collection | 4b. Type of Biospecimen | 4c. Type of Testing Conducted | 4d. Qualitative Results | 4e. Quantitative Result (e.g., viral load) | 4f. Specify units for Quantitative Results |
|---|---|---|---|---|---|---|
| SD1. | DD:MM:YY | • Amniotic fluid<br>• Placental swab (any side)<br>• Placental swab (fetal side)<br>• Cord blood<br>• Other, specify | • Viral PCR<br>• IgM<br>• IgG<br>• Other, specify | • Positive<br>• Negative<br>• Other, specify | ___ | ________<br>________ |
| SD2. | | | | | | |
| SD3. | | | | | | |

| SD4. |
|---|

#### Q6. Infant Biospecimen Table

| Sample Number (as needed) | 6a. Date of Biospecimen Collection | 6b. Type of Biospecimen | 6c. Type of Testing Conducted | 6d. Qualitative Results | 6e. Quantitative Result (e.g., viral load) | 6f. Specify units for Quantitative Results |
|---|---|---|---|---|---|---|
| SI1. | DD:MM:YY | • Nasopharyngeal swab<br>• Feces/rectal swab<br>• Gastric swab<br>• Neonatal peripheral blood<br>• Other, specify | • Viral PCR<br>• IgM<br>• IgG<br>• Other, specify | • Positive<br>• Negative<br>• Other, specify | ___ | ________<br>________ |
| SI2. | | | | | | |
| SI3. | | | | | | |
| SI4. | | | | | | |
