## Supplemental Table 3 for "Protocol for a sequential, prospective meta-analysis to describe coronavirus disease 2019 (COVID-19) in the pregnancy and postpartum periods"

**Description of participating studies**

Table 1-1: ANCOV Kenya

| **Study Site/Name** | ANCOV Kenya: Antenatal Care during COVID-19 |
| --- | --- |
| **Investigators** | Victor Akelo^1^, Beth A. Tippett Barr^1^, Dickens Onyango^2^, Sammy Khagayi^3^ |
| **Institutional Affiliations** | ^1^CDC Kenya, ^2^Kisumu County Department of Health; ^3^Kenya Medical Research Institute-Center for Global Health Research |
| **Study Design** | Prospective cohort study |
| **Participants** | Pregnant women both positive and negative for SARS-COV-2 |
| **Recruitment** | Community health care workers identified pregnant women and referred them to ANC where they were screened for covid. Covid screening also at labor & delivery. |
| **Sample Size** | 2,500 |
| **Timeline** | Recruitment began in August 2020 through December 2021 |
| **Outcomes** | Primary Outcome Measures: Adverse maternal outcomes |
| **Notes** | None |

Table 1-2: PRIORITY Study

| **Study Site/Name** | Pregnancy Coronavirus Outcomes Registry (PRIORITY) |
| --- | --- |
| **Investigators** | Valerie Flaherman^1^, Stephanie Gaw^1^, Yalda Afshar^2^, Vanessa Jacoby^1^ |
| **Institutional Affiliations** | ^1^Univesity of California, San Francisco; ^2^University of California, Los Angeles |
| **Study Design** | Nationwide registry/prospective cohort study |
| **Participants** | Pregnant or recently pregnant people (within 6 weeks) who have a suspected or confirmed COVID-19 diagnosis. |
| **Recruitment** | People were recruited from all clinical sites across the United States if they were under investigation for COVID-19 or received a COVID-19 diagnosis. Eligible participants were identified through voluntary physician referrals or self-referrals. All participants in PRIORITY were enrolled remotely through the UCSF Coordinating Center. |
| **Sample Size** | 1,260 people enrolled, 65% with confirmed COVID-19. |
| **Timeline** | Enrollment March 2020 through October 2020. Following up newborns through 6 and 12 months |
| **Outcomes** | Maternal outcomes: To evaluate the presentation, disease course, and clinical outcomes for pregnant people infected with COVID-19 compared with those that are COVID-19 negative. We will query participants on disease presentation, course of infection, treatments received, incidence and risk of hospitalization and/or ICU stay, and time to complete recovery.  Pediatric outcomes: To assess pediatric outcomes among infants born to people with COVID-19 compared with those that are COVID-19 negative. |
| **Notes** | More information at: <https://priority.ucsf.edu/researchers> |

Table 1-3: Neonatal Complications of Coronavirus Disease (COVID-19) (United Kingdom)

| **Study Site/Name** | Neonatal Complications of Coronavirus Disease (COVID-19) |
| --- | --- |
| **Investigators** | Christopher Gale^1^ and Jennifer J. Kurinczuk^2^ |
| **Institutional Affiliations** | ^1^Imperial College London, ^2^University of Oxford |
| **Study Design** | Prospective cohort study/National active surveillance |
| **Participants** | 1.Neonates COVID-19 positive: Neonatal COVID-19 in babies (<29 days old) in neonatal units, pediatric intensive care units and other in-patient locations.  2.Neonates born to COVID-19 positive mothers: Neonates (<29 days old) born to COVID-19 positive mothers requiring neonatal care |
| **Recruitment** | Pediatricians reported any neonate who met the case definition of neonatal complications coronavirus disease. |
| **Sample Size** | 500 |
| **Timeline** | March 2020 through March 2021 with a 6-month follow-up |
| **Outcomes** | Primary Outcome Measures:   1. Incidence of neonatal COVID-19 [ Time Frame: April 2020 to March 2021]   Number of neonatal participants with COVID-19 divided by the total number of live births in the population   2. Incidence of vertically transmitted COVID-19 [ Time Frame: April 2020 to March 2021]   Number of neonatal participants with COVID-19 following vertically transmission of the Coronavirus divided by the total number of live births in the population  Secondary Outcome Measures:   1. Presentation and natural history of neonatal COVID-19 [ Time Frame: April 2020 to March 2021] -Questionnaire data 2. Presentation of neonates with COVID-19 positive mothers [ Time Frame: April 2020 to March 2021] - Questionnaire data 3. Outcomes for neonates with COVID-19 [ Time Frame: April 2020 to March 2021] - Proportion of neonate participants who died and the proportion who were discharged home alive. 4. Clinical treatment of neonatal COVID-19 [ Time Frame: April 2020 to March 2021] - Questionnaire data 5. Neonatal secondary impacts of maternal COVID-19 [ Time Frame: April 2020 to March 2021] - Questionnaire data |
| **Notes** | These data reflect neonatal outcomes of pregnant cases identified by the UKOSS study.  Trial protocol: <https://clinicaltrials.gov/ct2/show/NCT04386109> |

Table 1-4: Madrid Hospital-Based Registry

| **Study Site/Name** | Madrid Hospital-based registry |
| --- | --- |
| **Investigators** | Mar Gil; Irene Fernandez Buhigas |
| **Institutional Affiliations** | Obstetrics and Gynecology, Hospital Universitario de Torrejón, Torrejón de Ardoz, Madrid, Spain. School of Medicine, Universidad Francisco de Vitoria, Pozuelo de Alarcón, Madrid, Spain |
| **Study Design** | Registry |
| **Participants** | Pregnant women with confirmed covid-19 diagnosis |
| **Recruitment** | Pregnant women screened for covid during antenatal care and labor and delivery, as well as if the patient presented with symptoms at any obstetric visit. |
| **Sample Size** | 200 cases as of April 2021 |
| **Timeline** | Ongoing |
| **Outcomes** | Primary Outcome Measures: effect of COVID-19 on the pregnancy outcomes and the effect of the pregnancy on the COVID-19 evolution. |
| **Notes** | None |

Table 1-5: Gestacovid: Registro Chileno de Embarazadas con Covid-19

| **Study Site/Name** | Gestacovid: Registro Chileno de Embarazadas con Covid-19 |
| --- | --- |
| **Investigators** | Jorge Carrillo^1^, Olivia Hernandez Bellolio^2^ |
| **Institutional Affiliations** | ^1^Departmento de Obstetricia y Ginecologia, Clinica Alemana de Santiago, Facultad de Medicina Clinica Alemana-Universidad del Desarrollo, Santiago, Chile  ^2^ Gynecology and Obstetrics, Felix Bulnes Hospital and RedSalud Clinic, Santiago, Chile |
| **Study Design** | National multicenter prospective cohort study |
| **Participants** | Active cases (pregnant and recently pregnant women) from various hospitals along Chilean territory); new sites may add controls |
| **Recruitment** | Covid screening done when patient was hospitalized for labor and delivery, during antenatal care visits, symptoms consulting, or patients who were in contact with someone positive. |
| **Sample Size** | 1,500 estimated recruitments |
| **Timeline** | Started in April 2020 and recruited last patients by August 31, 2020, with follow-ups completed in March 2021 |
| **Outcomes** | Natural history of neonatal COVID-19; maternal and perinatal outcomes. |
| **Notes** | None |

Table 1-6: Maternal and perinatal outcomes of pandemic influenza or novel coronavirus in pregnancy in the U.K. (UKOSS)

| **Study Site/Name** | Maternal and perinatal outcomes of pandemic influenza or novel coronavirus in pregnancy in the U.K. (UKOSS) |
| --- | --- |
| **Investigators** | Marian Knight |
| **Institutional Affiliations** | National Perinatal Epidemiology Unit, Nuffield Department of Population Health, University of Oxford |
| **Study Design** | National population-based cohort study using the UK Obstetric Surveillance System/International Network of Obstetric Survey Systems |
| **Participants** | All pregnant women in the UK admitted to hospital with confirmed novel coronavirus. |
| **Recruitment** | Cases will be identified through the UKOSS network of nominated reporting clinicians in each consultant led maternity unit in the UK (194 maternity units total). Nominated reporting clinicians will be asked to report all pregnant women with confirmed pandemic influenza or novel coronavirus admitted to their unit. |
| **Sample Size** | 1,253 cases as of September 2020. Expecting 7,000+. |
| **Timeline** | Interim results: August 2020 and published in September 2020. End date July 2021. |
| **Outcomes** | Primary Outcome Measures: Maternal death; Maternal level 3 critical care unit admission; Other major maternal complication; Preterm birth; Congenital anomaly; Perinatal death |
| **Notes** | More information at: <https://www.npeu.ox.ac.uk/ukoss/current-surveillance/covid-19-in-pregnancy> |

Table 1-7: COVID-19 Surveillance to Inform Public Health Action in Mali

| **Study Site/Name** | COVID-19 Surveillance to Inform Public Health Action in Mali |
| --- | --- |
| **Investigators** | Karen Kotloff^1^, Milagritos Tapia^1^, Amanda Driscoll^1^, Samba Sow^2^, Fadima Cheick Haidara^2^, Adama Mamby Keita^2^, Uma Onwuchekwa^2^ |
| **Institutional Affiliations** | ^1^University of Maryland, Baltimore  ^2^Centre pour le Dévelopement des Vaccins du Mali (CVD-Mali) |
| **Study Design** | Prospective cohort: mother-infant linked pregnancy surveillance |
| **Participants** | Covid positive and negative pregnant women |
| **Recruitment** | Cohorts: (see protocol for full inclusion/exclusion criteria)   1. Antenatal surveillance cohort (ANC-1): pregnant women recruited at ANC-1 visit 2. Antenatal surveillance cohort (DSS): pregnant women recruited from the community (DSS area) 3. Antenatal illness surveillance cohort: pregnant women who meet COVID-19 testing criteria and pregnant women who do not meet COVID-19 testing criteria |
| **Sample Size** | 1. ANC-1 pregnant woman cohort: 2,000 pregnant women and their infants 2. DSS pregnant people cohort: 1,000 pregnant women and their infants 3. ANC illness surveillance cohort: up to 1,500 pregnant women and their infants (up to 500 symptomatic + 1000 asymptomatic) |
| **Timeline** | From August 2020 through 6 weeks postpartum and 6 months postnatal for infants. The study is expected to be approximately 10 months but will depend on the on the trajectory of the COVID-19 outbreak in Mail. |
| **Outcomes** | Primary Outcomes: Mother-Infant linked Pregnancy Surveillance   - Life-threatening or potentially life-threatening maternal complications (e.g. stroke, hemorrhage, seizure, etc.) - maternal death - fetal loss prior to 22 weeks gestation - preterm birth (birth at <37 weeks gestation) - low birth weight (<2500g) - stillbirth - infant death within the first 6 months of life   Secondary outcomes: Pregnancy cohort:   - Maternal antenatal or postpartum requirement for hospitalization (excluding the delivery visit) - Infant requirement for hospitalization within the first six months of life - Congenital malformation at birth - SARS-CoV-2 microbiologically confirmed infection during pregnancy - SARS-CoV-2 microbiologically confirmed infection during the post-partum period - SARS-CoV-2 infection in the first six months of life - SARS-CoV-2 microbiologically confirmed infection in health care workers - SARS-CoV-2 seropositivity at delivery - SARS-CoV-2 seropositivity at the ANC-1 visit |
| **Notes** | The ANC-1 and DSS prospective surveillance cohorts are mutually exclusive but are designed to be analyzed together. |

Table 1-8: PERICOVID – PREPARE

| **Study Site/Name** | PERICOVID – PREPARE |
| --- | --- |
| **Investigators** | Kirsty Le Doare^1^, Gordon Rukundo^2^, Sammy Khagayi^3^ |
| **Institutional Affiliations** | ^1^ St. George’s, University of London; ^2^PeriCovid (PREPARE) – Uganda Team, Makerere University – Johns Hopkins University Research Collaboration, Uganda; ^3^ Kenya Medical Research Institute |
| **Study Design** | Prospective cohort study |
| **Participants** | Pregnant women at any gestation attending designated study centers for ANC and delivery |
| **Recruitment** | In 4 hospitals in Kampala, Uganda from ANC visits. Covid screening also at labor & delivery, as well as outpatient covid care and if patient presents with symptoms or exposure. |
| **Sample Size** | 25,000 (from Uganda and Malawi), 400 with covid symptoms |
| **Timeline** | 12 months (September 2020 to August 2021) |
| **Outcomes** | Primary Outcomes:   - The detection of an antibody against the spike protein of the SARS-CoV-2 virus in the serum of pregnant women - Infant colonization at day 0 - Pregnancy and neonatal outcomes: placental disorders, gestational hypertension, gestational diabetes, fetal growth restriction, all-cause maternal and neonatal mortality |
| **Notes** | More information at: <https://www.pericovid.com/pericovid-in-africa> |

Table 1-9: Washington State COVID-19 in Pregnancy Collaborative

| **Study Site/Name** | Washington State COVID-19 in Pregnancy Collaborative |
| --- | --- |
| **Investigators** | Kristina Adams Waldorf; Erica Lokken |
| **Institutional Affiliations** | University of Washington, Seattle, WA |
| **Study Design** | Retrospective medical records review of PCR confirmed SARS-CoV-2 infections in pregnancy (case-only) |
| **Participants** | Pregnant patients with PCR confirmed SARS-CoV-2 infections during any trimester of pregnancy detected through June 30, 2020 |
| **Recruitment** | Site-specific algorithms and medical records from 35 hospitals and clinics in Washington State. |
| **Sample Size** | 240 cases in pregnancy |
| **Timeline** | Data collection completed in September 2020. Data shared for PMA in February 2021. |
| **Outcomes** | Pregnancy, delivery, and some neonatal outcomes. COVID-19 disease severity and hospitalization data. |
| **Notes** | None |

Table 1-10: NICHD Global Network

| **Study Site/Name** | NICHD Global Network |
| --- | --- |
| **Investigators** | Beth McClure |
| **Institutional Affiliations** | RTI International (data coordinating center); ICDDR,B (Bangladesh); Moi University (Kenya); INCAP (Guatemala); Kinshasa School of Public Health (DRC); Lata Medical Research Foundation (India); KLE University (India); Aga Khan University (Pakistan); University of Zambia (Zambia); US Partners: University of Alabama at Birmingham; Boston University; Columbia University; University of Colorado; University of Virginia; University of North Carolina at Chapel Hill; Thomas Jefferson University; Indiana University |
| **Study Design** | Prospective cohort |
| **Participants** | All pregnancies within the catchment area are recruited. |
| **Recruitment** | Questionnaires used to identify suspected COVID-19, as well as ongoing antibody testing at delivery. A subset of people will also have antibody testing conducted at the first ANC visit. |
| **Sample Size** | Target of 2,500 pregnancies per study site (20,000 total) |
| **Timeline** | Expected 1-year timeline |
| **Outcomes** | Maternal morbidity and mortality; stillbirth; neonatal morbidity and mortality |
| **Notes** | None |

Table 1-11: NICHD Maternal-Fetal Medicine Units (MFMU) Network

| **Study Site/Name** | NICHD Maternal-Fetal Medicine Units (MFMU) Network |
| --- | --- |
| **Investigators** | Torri Metz^1^ (Protocol Chair), Rebecca Clifton^2^ (Data Coordinating Center PI) |
| **Institutional Affiliations** | ^1^University of Utah Health, ^2^ The George Washington University |
| **Study Design** | Registry and Observational cohort study |
| **Participants** | Pregnant people confirmed to have COVID-19 in participating network sites; the cohort component of the study includes random deliveries from 2020 at selected study sites for comparison. |
| **Recruitment** | Medical record abstraction following identification of a pregnant patient with a positive SARS-CoV-2 test at one of the sites in the MFMU. |
| **Sample Size** | Approximately 4,000 COVID-19 cases; the cohort study component will include approximately 10,000 random deliveries from the same study sites as unexposed. |
| **Timeline** | Recruitment began in June 2020 and medical record abstraction anticipated to be completed in July 2021 |
| **Outcomes** | The primary endpoint is a maternal composite defined as at least one of the following during pregnancy and through 6 weeks postpartum: mortality, morbidity related to hypertensive disorders of pregnancy, morbidity related to postpartum hemorrhage, morbidity related to infection. |
| **Notes** | More Information at: <https://mfmunetwork.bsc.gwu.edu/PublicBSC/MFMU/MFMUPublic/> |

Table 1-12: Canadian Surveillance of COVID-19 in Pregnancy

| **Study Site/Name** | Canadian Surveillance of COVID-19 in Pregnancy: Epidemiology, Maternal and Infant Outcomes (CANCOVID-Preg) |
| --- | --- |
| **Investigators** | Deborah Money |
| **Institutional Affiliations** | University of British Columbia |
| **Study Design** | National surveillance: prospective observational/surveillance cohort |
| **Participants** | People currently pregnant or delivered, living in the selected Canadian provinces (BC, AB, QC, and MB), documented SARS-CoV-2 infection in pregnancy |
| **Recruitment** | Identified by clinical or public health report of suspected COVID-19 and/confirmatory SARS-CoV-2 laboratory result. Care providers from the extensive reproductive infectious disease and maternity care network will identify potential participants. |
| **Sample Size** | 1,880 as of February 2021; varying sample sizes from each province |
| **Timeline** | April 2020-December 2023 |
| **Outcomes** | Primary Outcome Measures: Maternal outcomes will include the risk for preterm birth and delivery complications. Fetal outcomes will include Apgar scores at 1 and 5 minutes, birthweight, admission to NICU, positive test for SARS-CoV-2, and need for resuscitation at delivery |
| **Notes** | More information at: <https://ridprogram.med.ubc.ca/cancovid-preg/> |

Table 1-13: China and Hong Kong COVID Registry

| **Study Site/Name** | China and Hong Kong COVID Registry |
| --- | --- |
| **Investigators** | Liona C. Poon^1^; Huixia Yang^2^ |
| **Institutional Affiliations** | ^1^The Chinese University of Hong Kong; ^2^Peking University Health Science Center |
| **Study Design** | Registry |
| **Participants** | Pregnant women. Hong Kong: PCR positive. Original China data used probable infection definition, but new cases will be PCR. |
| **Recruitment** | Covid screening at labor and delivery. Clinical records were retrospectively reviewed from 25 hospitals in China, and cases recruited from all public hospitals across Hong Kong. |
| **Sample Size** | 146 ongoing case identification; China: 116. Hong Kong: 30. Following pregnant women and infants. |
| **Timeline** | Contribute initial data by August 2020. |
| **Outcomes** | Primary outcome measures: Maternal morbidity and mortality, birth outcomes, and neonatal mortality |
| **Notes** | None |

Table 1-14: RECOGEST Colombia

| **Study Site/Name** | RECOGEST Colombian registry: COVID and pregnancy |
| --- | --- |
| **Investigators** | Jose Sanin-Blair^1,7^, Nataly Velasquez^1^, Viviana Marcela Mesa Ramírez ^1,5^ , Jorge E. Tolosa^2,3,4^ , Yudy Alexandra Aguilar^1^, María Nazareth Campo Campo^1^ , Catalina Valencia ^8^ , Daniela Nasner^5^, Arturo Cardona Ospina^6^ , Mario Carvajal^8^ , Pablo Galvis ^9^, Maria Cristina Geney^10^, Jose Rojas-Suarez ^11,12^, Jose Santacruz^11^ , Beatriz Suarez^11^ , Juan Fernández ^11, 13^, Jezid Miranda^11^, Paula Velásquez^14^, Karina Ardila^15^, Marlene Baena^18 ,19,20^, Julio Duran^16,17^, Camilo Bello^11^, Walter Anicharico^11^, Diana Borre^11^ |
| **Institutional Affiliations** | ^1^Clinica Universitaria Bolivariana, ^2^ FUNDARED-MATERNA , ^3^St. Luke’s University Health Network, ^4^ Oregon Health & Science University, ^5^ Fundación Valle del Lili, ^6^ Clinica del Prado,^7^Medicina Fetal SAS.^8^Clinica Somer, ^9^ Norfetus, ^10^ Hospital Universitario Clinica San Rafael. ^11^Intensive Care and Obstetrics Research Group (GRICIO), Universidad de Cartagena, ^12^Corporación Universitaria Rafael Núñez, ^13^EPS Mutual Ser, ^14^Clinica Versalles, ^15^Universidad libre Sección Cali, ^16^Instituto Cardiovascular del Cesar, ^17^Clinica Integral de Emergencia Laura Daniela, ^18^ ESE Hospital san jeronimo, ^19^ Clinica evaluamos , ^20^ Clinica materno infantil casa del niño . |
| **Study Design** | Multicenter National Registry: prospective, longitudinal observational study |
| **Participants** | All COVID-19 positive pregnant and puerperal women in the participating institutions. |
| **Recruitment** | Participants identified in 9 hospitals and 2 ambulatory settings. |
| **Sample Size** | 60 cases as of April 2021, expected to be 500 patients total. |
| **Timeline** | Recruitment ended December 2020 |
| **Outcomes** | Primary Outcome Measures: Maternal, perinatal, and fetal outcomes and socioeconomic characteristics. Emphasis in COVID positive admitted to Intensive Care Unit |
| **Notes** | More information at: <https://www.clinicauniversitariabolivariana.org.co/clinica/es/recogest?resolvetemplatefordevice=true> |

Table 1-15: PERICOVID Africa – PRECISE

| **Study Site/Name** | PERICOVID – PRECISE |
| --- | --- |
| **Investigators** | Peter von Dadelszen |
| **Institutional Affiliations** | King’s College London |
| **Study Design** | Prospective cohort study |
| **Participants** | Pregnant women at any gestation attending designated study centers for ANC and delivery in the Gambia, Kenya, and Mozambique. |
| **Recruitment** |  |
| **Sample Size** | Tentative target: 6,000 |
| **Timeline** | Tentative: August 2020 through June 2021 |
| **Outcomes** | Key outcomes include:   - Stillbirth - Pre-term birth - Hypertension |
| **Notes** | More information at: <https://www.pericovid.com/pericovid-in-africa> |

Table 1-16: CHOPAN Registry

| **Study Site/Name** | Coronavirus Health Outcomes in Pregnancy and Newborns (CHOPAN) Registry |
| --- | --- |
| **Investigators** | Clare Whitehead |
| **Institutional Affiliations** | University of Melbourne |
| **Study Design** | Registry of pregnant people who are infect with COVID-19 in Australia and New Zealand |
| **Participants** | Pregnant people with confirmed COVID-19 (suspected cases are not included) |
| **Recruitment** | Pregnant people admitted to a participating hospital. |
| **Sample Size** | 50 cases as of August 2020 (targeting 200 cases) |
| **Timeline** | Enrollment occurred between April 2020 and April 2021. Date of last data collection: December 2021. |
| **Outcomes** | Obstetric, perinatal, and neonatal outcomes after coronavirus infection |
| **Notes** | CHOPAN is collaborating with EpiCentre on neonatal outcomes. More information at: <https://www.anzctr.org.au/Trial/Registration/TrialReview.aspx?id=379572&isReview=true> |

Table 1-17: Puerto Rico CASSS

| **Study Site/ Name** | Puerto Rico COVID-19 Antenatal Sentinel Surveillance System (CASSS) |
| --- | --- |
| **Investigators** | Miguel Valencia-Prado^1^, Camille Delgado^2^ |
| **Institutional Affiliations** | ^1^Children with Special Medical Needs Division, Puerto Rico Department of Health, San Juan, Puerto Rico; ^2^Surveillance for Emerging Threats to Mothers and Babies, Puerto Rico Department of Health, San Juan Puerto Rico |
| **Study Design** | Active Surveillance |
| **Participants** | Confirmed covid positive pregnant people on the island. |
| **Recruitment** | New cases identified: Received daily reports of confirmed and suspected pregnant people cases from the Puerto Rican Department of Health BioPortal regional and municipal epidemiologists, and SARS-CoV-2 Contact Tracing Group. Cases also identified by Ob-Gyns in participating, and other medical staff on all medical facilities on the island and reported directly. Also recruited through monthly data linkages where birth certificates database was compared with the Puerto Rico Department of Health SARS-CoV-2 Case Surveillance System database to identify new cases. |
| **Sample Size** | Monitoring 458 cases as of March 2021 |
| **Timeline** | Ongoing |
| **Outcomes** | Maternal mortality and morbidity, birth outcomes, and neonatal mortality |
| **Notes** | None |

Table 1-18: AFREHealth

| **Study Site/Name** | The African Forum for Research and Education in Health’s COVID-19 Research Working Group (AFREHealth) |
| --- | --- |
| **Investigators** | Jean B. Nachega^1-4^; Nadia A. Sam-Agudu^5,6,7^; Catherine Cluver^8,9^; Adrie Bekker^10^; Liesl de Waard^8^; Onesmus W. Gachuno^11^; John Kinuthia^11,12^; Nancy Mwongeli^12^; Samantha Budhram^13^; Valerie Vannevel^14^; Priya Somapillay^15^; Elizabeth Agyare^16^; Akwasi Baafuor Opoku^17^; Aminatu Umar Makarfi^18^; Asara M. Abdullahi^19^; Daniel Katuashi Ishoso^20^; Michel Tshiasuma Pipo^21^; Christian Bongo-Pasi Nswe,^21,22^; John Ditekemena^20^; Ethan Borre^23^; Lovemore Nyasha Sigwadhi^24^; Rhoderick N. Machekano^24^; Peter S. Nyasulu^24^; Michel P. Hermans^25^; Musa Sekikubo^26^; Christopher Nsereko^27^; Philippa Musoke^26^; Evans K. Agbeno^28^; Michael Yaw Yeboah^17^; Lawal W. Umar^29^; Mukanire Ntakwinja^30^; Denis M. Mukwege^30^; Etienne Kajibwami Birindwa^31^; Serge Zigabe Mushamuka^31^; Emily R. Smith^32^; Eduard J. Mills^33^; John Otshudiema Otokoye^34^; Placide Mbala-Kingebeni^35^; Jean-Marie N. Kayembe^36^; Don Jethro Mavungu Landu^19,20^; Jean-Jacques Muyembe Tamfum^35^; Alimuddin Zumla^37,38^; Aster Tsegaye^39^; Alfred Mteta^40^; Nelson Sewankambo^41^; Fatima Suleman^42^; Prisca Adejumo^43^; Jean R. Anderson^44^; Emilia V. Noormahomed^45^; Richard J. Deckelbaum^46^; Jeff Stringer^47^; Abdon Mukalay^48^; Taha E. Taha^3^; Mary Glenn-Fowler^49^; Judith N. Wasserheit^50^; Refiloe Masekela^51^; Lynne M. Mofenson^52^; Eduard Langenegger^8^ |
| **Institutional Affiliations** | 1. Department of Medicine, Division of Infectious Diseases, Stellenbosch University Faculty of Medicine and Health Sciences, Cape Town, South Africa 2. Department of Epidemiology, Infectious Di0seases and Microbiology, and Center for Global   Health, University of Pittsburgh, Pittsburgh, PA, USA   3. Department of Epidemiology, Johns Hopkins Bloomberg School of Public Health, Baltimore, MD, USA 4. Department of International Health, Johns Hopkins University, Bloomberg School of   Public Health, Baltimore, MD, USA   5. International Research Center of Excellence, Institute of Human Virology Nigeria, Abuja, Nigeria 6. Department of Paediatrics and Child Health, University of Cape Coast School of Medical Sciences, Cape Coast, Ghana 7. Institute of Human Virology and Department of Pediatrics, University of Maryland School of Medicine, Baltimore, MD, USA 8. Department of Obstetrics and Gynecology; Faculty of Medicine and Health Sciences, Stellenbosch University, Cape Town, South Africa 9. Translational Obstetrics Group and Mercy Perinatal, Department of Obstetrics and Gynaecology, University of Melbourne, Victoria, Australia 10. Department of Paediatrics and Child Health; Faculty of Medicine and Health Sciences, Stellenbosch University, Cape Town, South Africa 11. Department of Obstetrics and Gynaecology, University of Nairobi, Nairobi, Kenya 12. Department of Research, Department of Reproductive Health, Kenyatta National Hospital, Nairobi, Kenya 13. Department of Obstetrics and Gynecology, University of KwaZulu Natal, Durban, South Africa 14. Department of Obstetrics and Gynecology, Kalafong Hospital, University of Pretoria, Pretoria, South Africa 15. Maternal Foetal Medicine; Steve Biko Hospital, University of Pretoria, Pretoria, South Africa 16. Department of Microbiology, School of Medical Sciences, University of Cape Coast and Cape Coast Teaching Hospital, Cape Coast, Ghana 17. Department of Obstetrics and Gynaecology, Komfo Anokye Teaching Hospital, Kumasi, Ghana 18. Department of Obstetrics and Gynaecology, College of Health Sciences, Ahmadu Bello University and Ahmadu Bello University Teaching Hospital, Zaria, Nigeria 19. Department of Medicine, College of Health Sciences, Ahmadu Bello University and Ahmadu Bello University Teaching Hospital, Zaria, Nigeria 20. Department of Community Health, School of Public Health, University of Kinshasa, Kinshasa, Democratic Republic of the Congo 21. Faculty of Public Health, Université Moderne de Kinkole, Kinshasa, Democratic Republic of the Congo 22. Department of Public Health, Centre Interdisciplinaire de Recherche en Ethnopharmacologie, Faculty of Medicine, Université Notre-Dame du Kasayi, Kananga, Democratic Republic of the Congo 23. Department of Medicine, Duke University School of Medicine, Durham, NC, USA 24. Division of Epidemiology and Biostatistics, Department of Global Health, Faculty of Medicine and Health Sciences, Stellenbosch University, Cape Town, South Africa 25. Department of Endocrinology and Nutrition, Cliniques Universitaires St-Luc, Brussels, Belgium 26. Dept of Paediatrics & Child Health, School of Medicine, Makerere University, Kampala, Uganda 27. Department of Medicine, Entebbe Regional Reference Hospital, Entebbe, Uganda 28. Department of Obstetrics & Gynecology, School of Medical Sciences, University of Cape Coast and Cape Coast Teaching Hospital, Cape Coast, Ghana 29. Department of Pediatrics, College of Health Sciences, Ahmadu Bello University and Ahmadu Bello Teaching Hospital, Zaria, Nigeria 30. Gynaecology and General Surgery, Panzi General Referral Hospital, Bukavu, Democratic Republic of the Congo 31. Hôpital Provincial Général de Référence de Bukavu and Faculty of Medicine, Université Catholique de Bukavu, Bukavu, Democratic Republic of the Congo 32. Department of Global Health, Milken Institute School of Public Health, The George Washington University, Washington DC, USA 33. Department of Health Research Evidence and Impact, Faculty of Health Sciences, McMaster University, Hamilton, Canada 34. Epidemiological Surveillance Team, COVID-19 Response, Health Emergencies Program, World Health Organization, Kinshasa, Democratic Republic of the Congo 35. Department of Medical Microbiology and Virology, Faculty of Medicine, University of Kinshasa, National Institute of Biomedical Research , Kinshasa, Democratic Republic of the Congo 36. Department of Medicine, Faculty of Medicine, University of Kinshasa, Kinshasa, Democratic Republic of the Congo 37. Division of Infection and Immunity, Department of Infection, Centre for Clinical Microbiology, University College London, London, United Kingdom 38. National Institute for Health Research Biomedical Research Centre, University College London Hospitals, London, United Kingdom 39. Department of Medical Laboratory Sciences, College of Health Sciences, Addis Ababa University, Addis Ababa, Ethiopia 40. Kilimanjaro Christian Medical University College, Moshi, United Republic of Tanzania 41. School of Medicine, College of Health Sciences, Makerere University, Kampala, Uganda 42. Discipline of Pharmaceutical Sciences, University of KwaZulu-Natal, Durban, South Africa 43. Department of Nursing, University of Ibadan, Ibadan, Nigeria 44. Department of Obstetrics and Gynecology, Johns Hopkins School of Medicine, Baltimore, MD, USA 45. Faculty of Medicine, Eduardo Mondlane University, Maputo, Mozambique 46. Department of Pediatrics, Columbia University Irving Medical Center, New York, NY, USA 47. Department of Obstetrics and Gynecology, University of North Carolina, School of Medicine, Chapel Hill, NC, USA 48. Faculty of Medicine, University of Lubumbashi, Lubumbashi, Democratic Republic of the Congo. 49. Department of Pathology, Johns Hopkins University School of Medicine, Baltimore, MD, USA 50. Departments of Global Health and Medicine, Schools of Medicine and Public Health, University of Washington, Seattle, WA, USA 51. Department of Pediatrics and Child Health, School of Clinical Medicine, College of Health Sciences, University of KwaZulu Natal, Durban, South Africa 52. Elizabeth Glaser Pediatric AIDS Foundation, Washington DC, USA |
| **Study Design** | Retrospective cohort of pregnant women who are infected with COVID-19 in the DRC, Ghana, Kenya, Nigeria, South Africa, and Uganda. |
| **Participants** | Pregnant women with confirmed COVID-19; non-COVID pregnant women and non-pregnant COVID positive women (comparison groups) |
| **Recruitment** | Retrospective medical records/charts review |
| **Sample Size** | 350 ongoing case identification. Varying sample sizes in each country. |
| **Timeline** | March 2020 to October 2020 |
| **Outcomes** | Maternal outcomes, maternal death, and neonatal outcomes |
| **Notes** | None |

Table 1-19: Covi-Preg

| **Study Site/Name** | Covi-Preg |
| --- | --- |
| **Investigators** | Alice Panchaud^1, 2^ ; Guillaume Favre^3^ |
| **Institutional Affiliations** | ^1^Institute of Primary Health Care (BIHAM), University of Bern, Bern, Switzerland ^2^Service of Pharmacy, Lausanne University Hospital and University of Lausanne, Lausanne Switzerland  ^3^ Materno-fetal and Obstetrics Research Unit, Department “Femme-Mère-Enfant”, University Hospital, Lausanne, Switzerland |
| **Study Design** | International Registry; cohort study |
| **Participants** | Pregnant women with suspected or confirmed covid cases in >150 hospitals worldwide. |
| **Recruitment** | An international network that collected data from the participating institutions. Universal covid screening done at labor & delivery, as well as any outpatient visit or testing site when patient presented with symptoms. |
| **Sample Size** | 1,700 cases total. Varying sample sizes in each participating country. |
| **Timeline** | Enrollment began in March 2020. Date of planned final study report: July 2023. |
| **Outcomes** | Maternal mortality and morbidity, birth outcomes, and neonatal mortality |
| **Notes** | None |

Table 1-20: Fondazione Policlinico Universitario Agostino Gemelli, IRCCS, Rome

| **Study Site/Name** | Fondazione Policlinico Universitario Agostino Gemelli, IRCCS, Rome |
| --- | --- |
| **Investigators** | Elisa Bevilacqua^1^, Valentina Laurita Longo^2^, Federica Meli^2^, Giulia Bonanni^2^, Antonio Lanzone^1, 2^ |
| **Institutional Affiliations** | ^1^Department of Women and Child Health, Women Health Area, Fondazione Policlinico Universitario Agostino Gemelli, IRCCS, Rome, Italy  ^2^Catholic University of Sacred Hearth, Rome, Italy |
| **Study Design** | Prospective Cohort/registry |
| **Recruitment** | Covid screening at labor & delivery, as well as if patient presents with symptoms or exposure. |
| **Participants** | All pregnant women included (covid and non-covid) |
| **Sample Size** | Ongoing case identification: 230 |
| **Timeline** | PregOutCOV study: February to November 2020 |
| **Outcomes** | PregOutCOV Study outcomes include: composite obstetrical and neonatal adverse outcomes |
| **Notes** | The first 109 cases are included in the following study: *Pregnancy Outcomes according to the gestational age of acquiring COVID-19: an international case-control study (PregOutCOVstudy).* There are currently 3184 controls (non-covid pregnancies) 8 cases were included in reports from the World Association of Perinatal Medicine working group on COVID-19. The remaining individual patient data have not been included in any current study. |

Table 1-21: ARTIST India (FOGSI)

| **Study Site/Name** | Asian Research and Training Institute for Skill Transfer (ARTIST) India |
| --- | --- |
| **Investigators** | Hema Divakar |
| **Institutional Affiliations** | Asian Research and Training Institute for Skill Transfer (ARTIST), Bengaluru, India |
| **Study Design** | Registry |
| **Participants** | Pregnant women with confirmed COVID-19 cases in two major study centers in Karnataka. |
| **Recruitment** | The patients were screened when they came for ANC visits or for delivery for suspected covid positive. The patients were advised to get a test. If the test report is positive for COVID-19, the participated were recruited for the study. |
| **Sample Size** | 214 cases sent to date. Expecting 300 case identifications. |
| **Timeline** | Ongoing |
| **Outcomes** | Maternal mortality and morbidity, birth outcomes, and neonatal mortality |
| **Notes** | Study site is also contributing to Pan-COVID |

Table 1-22. Mexico National Registry

| **Study Site/Name** | Mexico National Registry |
| --- | --- |
| **Investigators** | Raigam Jafet Martinez-Portilla |
| **Institutional Affiliations** | National Institute of Perinatology, Mexico City, Mexico |
| **Study Design** | National Registry/population based-surveillance |
| **Participants** | Pregnant and non-pregnant women admitted to any of the 475 participating hospitals with a suspected COVID diagnosis. |
| **Recruitment** | Women who were admitted to any of the participating facilities for a suspected COVID diagnosis were tested based on clinical discretion. All tested patients were entered into the registry before testing, regardless of rt-PCR results, but the analysis was restricted to only positive patients. |
| **Sample Size** | 11,031 cases identified as of April 2021. |
| **Timeline** | February 2020 to present |
| **Outcomes** | Maternal mortality and morbidity, birth outcomes, and neonatal mortality |
| **Notes** | None |

Table 1-23: WHO Pregnancy and COVID-19 Cohort Study

| **Study Site/Name** | WHO Pregnancy and COVID-19 Cohort |
| --- | --- |
| **Investigators** | Nathalie Broutet, Anna Thorson (Responsible Officers), and the WHO Generic Protocol Implementation Team |
| **Institutional Affiliations** | World Health Organization (participating country investigators TBC) |
| **Study Design** | Prospective Cohort Study |
| **Participants** | Pregnant women (up to 2 days postpartum/post-pregnancy termination at time of enrollment) |
| **Recruitment** | Consecutive prospective recruitment will occur in health-care facilities, antenatal care clinics, COVID-19 testing centers, or at the community level. Pregnant minors will be offered inclusion in the study.  All pregnant women irrespective of SARS-CoV-2 testing results will be offered participation in the study until the sample sizes of exposed and unexposed groups are fulfilled. Recruitment timing and strategy in relation to pregnancy assessment and test/screening results may be implemented differently depending on the facilities and staffing resources at each study site.  All women will undergo virologic (RT-PCR or antigen-based) and serologic testing at enrollment.  Exposed (SARS-CoV-2 positive) pregnant women: Confirmed by positive RT-PCR or antigen or serologic test at enrolment or by documented positive test during pregnancy before enrollment. people  Unexposed (SARS-CoV-2 negative) women : Confirmed by negative virologic (RT-PCR or antigen) and serologic testing at enrollment:  Information will also be collected from all women about receipt of any doses of a COVID-19 vaccine; vaccinated women will be enrolled concurrently. |
| **Sample Size** | Varying sample sizes in each participating country according to national epidemic contexts and estimates of adverse composite outcomes. Current anticipated sample size for participating countries is >16,000. |
| **Timeline** | Anticipated May/June 2021 through Dec 2022; May vary by individual study site |
| **Outcomes** | Primary objective 1: To determine if SARS-CoV-2 infection in pregnancy increases the risk of adverse:   1. Maternal and pregnancy outcomes: (e.g. induced abortion, ectopic pregnancy, miscarriage, delivery mode, preterm birth, fetal growth restriction, haemorrhage, maternal morbidity and mortality); 2. Perinatal outcomes: (e.g. stillbirth, admission to the neonatal ICU, neonatal mortality within 7 days of life); 3. Neonatal outcomes: (e.g. morbidity, infection, mortality up to 4 weeks of life); 4. Postpartum outcomes: (e.g. infection, bleeding, transmission of SARS-CoV-2 up to 6 weeks after delivery).   Primary objective 2: To estimate the risk of MTCT of SARS-CoV-2 during pregnancy, intrapartum, postpartum or during breastfeeding among mother–neonate pairs with confirmed SARS-CoV-2 infection in pregnancy.  Primary objective 3: To evaluate clinical, pregnancy, and neonatal outcomes (as above) among women who receive at least one dose of COVID-19 vaccine during pregnancy. |
| **Notes** | The original generic protocol is available at: : <https://www.who.int/publications/m/item/a-prospective-cohort-study-investigating-maternal-pregnancy-and-neonatal-outcomes-for-women-and-neonates-infected-with-sars-cov-2> .  This study description also reflects amendments to the protocol which are currently undergoing ethics review. The updated protocol will be posted upon completion. |

Table 1-24: Pregnancy and Neonatal outcomes for women with COVID-19

| **Study Site/Name** | Pregnancy And Neonatal outcomes for women with COVID-19 (PAN-COVID) |
| --- | --- |
| **Investigators** | Edward Mullins^1^, Christoph Lees^1^, Julia Townson^2^, Rebecca Playle^2^ |
| **Institutional Affiliations** | ^1^Department of Metabolism, Digestion, and Reproduction, Imperial College London, United Kingdom; ^2^Center for Trials Research, Cardiff University, Wales, United Kingdom |
| **Study Design** | Registry |
| **Participants** | COVID positive and suspected COVID pregnant women |
| **Recruitment** | Universal covid screening at labor and delivery |
| **Sample Size** | N=8263 |
| **Timeline** | January 2020 – March 31, 2021 |
| **Outcomes** | Birth and neonatal outcomes |
| **Notes** | More information at: <https://pan-covid.org/> |

Table 1-25: Cerner Real World Data

| **Study Site/Name** | George Washington University / Adverse Perinatal Outcomes During The COVID-19 Pandemic |
| --- | --- |
| **Investigators** | Ethan A. Litman; Homa K. Ahmadzia |
| **Institutional Affiliations** | Department of Obstetrics & Gynecology, George Washington University, Washington, District of Columbia |
| **Study Design** | Retrospective cohort study |
| **Participants** | Covid positive and negative pregnant women |
| **Recruitment** | Identified via Labor and Delivery admission records during the study time period |
| **Sample Size** | 5,000 COVID positive pregnancies (260000 total sample) |
| **Timeline** | January 2020 – March 2021 |
| **Outcomes** | - COVID Positive PCR result; - Maternal mortality; - Prematurity; - Stillbirth (gestational age greater than 20 weeks); - Hypertensive disorders of pregnancy, eclampsia, HELLP; - Diabetes; - Postpartum hemorrhage; - Blood transfusion; - Preterm rupture of membranes; - Disseminated intravascular coagulation; - Myocardial infarction, cardiac arrest, heart failure; - Mechanical ventilation; - Aneurysm; - Acute Respiratory Distress Syndrome; - Amniotic fluid embolism; - Pulmonary edema; - Sepsis, shock; - Venous thromboembolism, arterial thromboembolism, cerebrovascular disease - Smoking; - Obesity; - Severe mental illness; - Cesarean Rate; - Ectopic pregnancy; - Birth weight; - 1 min Apgar; - 5 min Apgar |
| **Notes** | Cerner Real-World Data is extracted from the EMR of hospitals in which Cerner has a data use agreement. Encounters may include pharmacy, clinical and microbiology laboratory, admission, and billing information from affiliated patient care locations. All admissions, medication orders and dispensing, laboratory orders and specimens are date and time stamped, providing a temporal relationship between treatment patterns and clinical information. Cerner Corporation has established Health Insurance Portability and Accountability Act-compliant operating policies to establish de-identification for Cerner Real-World |

**Key stakeholder contributors**

| **Organization** | **Steering Committee Members** |
| --- | --- |
| American College of Obstetrics and Gynecology (ACOG) | Maria Diaz, Sara Homayouni, Nadia Ramey, Alireza A. Shamshirsaz |
| International Federation of Gynecology and Obstetrics (FIGO) | Jeanne Conry, Lesley Regan |
| Eunice Kennedy Shriver National Institute of Child Health and Human Development (NICHD) | Daniel Raiten, Nahida Chakhtoura, Caroline Signore |
| The World Health Organization (WHO) | WHO Generic Protocol Implementation Team (Ibukun Abejirinde, Edna Kara, Sami Gottlieb, Christine Godwin, Soe Soe Thwin, Mohamed Ali, Ndema Habib, Daniel Giordano), Rajiv Bahl, Mercedes Bonet Semenas, Karen M. Edmond, Caron Rahn Kim |
